# Skeletal muscle biomarkers of amyotrophic lateral sclerosis: a large-scale, multi-cohort proteomic study

**DOI:** 10.1101/2025.04.23.25326161

**Authors:** Oleksandr Dergai, Joanne Wuu, Magdalena Koziczak-Holbro, Andrea Malaspina, Volkan Granit, Jessica P. Hernandez, Anne Cooley, Ruchika Sachdev, Lili Yu, Michael Bidinosti, Ludvine Flotte, Mark Nash, Lori L. Jennings, James D. Berry, Lucie I Bruijn, Sophie Brachat, Michael Benatar

**Affiliations:** Novartis Diseases of Aging and Regenerative Medicine, Novartis Biomedical Research, CH-4002 Basel, Switzerland; Department of Neurology and ALS Center, University of Miami Miller School of Medicine, Miami, FL, USA; UCL Queen Square Motor Neuron Disease Center, UCL Queen Square Institute of Neurology, University College London, Queen Square, London, UK; Novartis Disease Area X, Novartis Biomedical Research, Cambridge, MA 02139, USA; Sean M. Healey and AMG Center for ALS, Massachusetts General Hospital, Boston, MA, USA; Biomarker Development, Novartis Pharmaceuticals, UK

**Keywords:** context of use, disease state biomarkers, monitoring biomarkers, disease progression biomarkers, pharmacodynamic response biomarkers

## Abstract

**Background:** Biomarkers with clear contexts-of-use are important tools for ALS therapy development. Understanding their longitudinal trajectory in the untreated state is key to their use as potential markers of pharmacodynamic response. To this end, we undertook a large-scale proteomic study in well-phenotyped cohorts to identify biomarker candidates of ALS disease state and disease progression.

**Methods:** Clinical phenotypic data and biofluid samples, collected from patients with ALS and healthy controls through multiple longitudinal natural history studies, were used to identify biomarker candidates. SOMAmer (Slow Off-rate Modified Aptamer)-based relatively quantitative measurement of ∼7,000 proteins was performed in plasma and CSF, with immunoassay validation of candidates of interest.

**Results:** We identified 329 plasma proteins significantly differentially regulated between ALS and controls (adjusted p-value <0.05), with 25 showing >40% relative abundance. PDLIM3, TNNT2, and MYL11 had the greatest log-fold elevation, while ANTXR2 and ART3 had the greatest log-fold reduction. A similar set of plasma proteins was found to increase (e.g. PDLIM3, TNNT2, MYL11) or decrease (e.g. ANTXR2, ART3, MSTN) with disease progression. CSF proteins with the greatest log-fold elevation included NEFL, NEFH, CHIT1, CA3, MYL11 and GPNMB. These results were confirmed in an independent replication cohort. Moreover, tissue-specific signature enrichment suggests a significant contribution of muscle as a source of these biomarkers. Immunoassays provided orthogonal validation of plasma TNNT2 and CSF GPNMB.

**Conclusion:** We identified an array of novel biomarkers with the potential to serve as response biomarkers to aid therapy development, as well as to shed light on the underlying biology of disease.

**(Key messages):** *What is already known on this topic:*

- There are currently few monitoring and disease progression biomarkers in ALS; and there is no published work from large-scale, multi-cohort proteomic studies that utilized longitudinal plasma and CSF samples to help fill this gap.

*What this study adds:*

- Using Slow Off-rate Modified Aptamer (SOMAmer)-based methods, we have identified an array of novel biomarkers of disease state (i.e. differentially regulated in ALS vs. controls) and ALS disease progression. These included, among others, PDLIM3, MYL11, ANTXR2, ART3, and MSTN.
- Skeletal muscle is the likely source of many of these newly discovered biomarkers.

*How this study might affect research, practice or policy:*

- These newly identified monitoring and disease progression biomarkers may be used to evaluate pharmacodynamic response in future clinical trials, thereby aiding ALS therapy development efforts.

## INTRODUCTION

Amyotrophic Lateral Sclerosis (ALS) is neurodegenerative disorder with very few therapeutics meaningfully impacting the course of disease.^1–3^ Biomarkers are increasingly recognized as essential tools to advance ALS therapy development, and their utility is critically dependent on well-defined contexts of use.^4^ In accordance with the FDA’s BEST Resource, biomarkers assessed repeatedly over time are *monitoring biomarkers*.^5^ Monitoring biomarkers that are dynamic (i.e. change over time) may be *markers of disease progression*. In ALS, urinary p75 neurotrophin receptor extracellular domain (p75^ECD^) is one such example.^6^ Monitoring markers that are relatively stable over time may be considered *temporally stable markers*, a concept (even if not a term) that is widely recognized.^7–9^ Neurofilament light chain (NfL)^10–12^ is one such example.^10–13^ Both types of monitoring biomarkers^13^ may be used to evaluate a potential pharmacodynamic response to therapeutic intervention.^3^ p75^ECD^ and NfL, however, each have their own limitations.^12^ Analytic variability of p75^ECD^ has limited its broader uptake, and changes in NfL concentration in response to a therapeutic may be confounded by short-term toxicity (e.g. AAV effect on dorsal root ganglia^14–16^) or an effect on NfL clearance.^17^ With increasing recognition of the essential role for biomarkers in phase 2 trials, ^18^ there is a pressing need to identify additional biomarkers with utility in monitoring and demonstrating response to an experimental therapeutic.

Here, we describe the use of the SomaScan platform for discovery and replication of both disease state (i.e. ALS vs. control) and monitoring (both disease progression and temporally stable) biomarker candidates in plasma and CSF from two independent longitudinal ALS (and healthy control) cohorts. We use the term “disease state” rather than “diagnostic biomarker” because the latter should entail discrimination between those with ALS and those with similarly appearing conditions at a time when the diagnosis is unclear^4^. In this manuscript, we focus on biomarkers relevant to therapy development for clinically manifest ALS (eFigure 1).

## MATERIALS AND METHODS

### Discovery and replication cohorts

Biofluid samples, with accompanying phenotypic data, for the discovery cohort were obtained from the University of Miami *Clinical Research in ALS (CRiALS)* study (clinicaltrials.gov NCT00136500) and the Clinical Research in ALS and Related Disorders for Therapeutic Development (CReATe) Consortium multi-center *Phenotype-Genotype-Biomarker (PGB1)* (NCT02327845) and *TRIAL READY* (NCT03912987) studies. For PGB1 and TRIAL READY, only samples and data collected at the Miami site were included. All 3 studies in the discovery cohort were approved by the University of Miami Institutional Review Board (IRB), which also serves as the single IRB of record for the CReATe Consortium, and all study participants provided written informed consent. For the replication cohort, biofluid samples and accompanying clinical information were obtained from 2 biomarker studies (NCT01495390; NCT00677768) through the Northeast ALS (NEALS) Biofluid Repository (https://neals-biorepository.smapply.org). Additional plasma and CSF samples from healthy controls were obtained from the Novartis Basel Tissue Donor Program (plasma, n=10) and the commercial National BioService, LLC, Saint-Petersburg, Russia (plasma, n=3; CSF, n=13). Samples used in the replication cohort were collected under the auspice of ethics/institutional review board approval, with written informed consent obtained from all study participants. The sample and data use in this project was approved by the responsible local ethics committee in Switzerland. Additional details are in the eMethods.

### Sample collection and processing

Biological specimens were collected, processed, and stored according to strict standard operating procedures, comparable across all studies, apart from the National BioService, for which limited information about processing times was available. Briefly, for plasma, blood was collected in K2 EDTA tubes, centrifuged at 1,750g for 10 minutes at 4°C within 2 hours of collection, and aliquoted for storage at −80°C. CSF was collected in polypropylene tubes (or polyethylene tubes for sample samples in the replication cohort), centrifuged (1,750g for 10 minutes at 4°C), aliquoted into polypropylene cryogenic sterile freestanding conical microtubes, frozen within ∼60 minutes of collection, and stored at −80°C.

### Proteomics measurements

The high-throughput SomaScan® platform was used to quantify relative levels of proteins in plasma and CSF samples. The SomaScan platform is based on modified single-stranded DNA aptamers (SOMAmer, Slow Off-rate Modified Aptamer) binding to specific protein targets with high specificity and sensitivity. The SomaScan Assay v4.1 used in this study measures 7,335 unique SOMAmers with a median intra-run and inter-run coefficients of variation (CV) of ∼5%.^19^ SomaScan analysis was performed at SomaLogic Inc. (Boulder, United States), with proteins quantified on Agilent hybridization arrays (Agilent Technologies) as previously described.^20,21^ Plasma samples were measured at 3 dilutions (20%, 0.5%, 0.005%) and CSF samples were measured at a single dilution (20%). Plasma and CSF samples were randomized across plates by disease status, sex and genotype to achieve a balanced plate layout and avoid technical batch effects driven by plate; samples from the same person were included on the same plate

### Statistical analysis

Raw intensity data was processed through the Somalogic pipeline with ANML normalization, quality control (QC), and outlier removal. Analyses were conducted separately for plasma and CSF, as protein quantification varied significantly between them. Exploratory analysis identified data structure and important covariates. Mixed effect linear models were employed for primary analyses to assess protein associations with covariates such as disease status and duration. Identified proteins underwent functional and expression enrichment analyses. Detailed QC and analytical steps are available in the eMethods.

### Immunoassay validation

TNNT2 (troponin T2) concentration was quantified using the Roche Elecsys Troponin T high sensitivity (hs) electrochemiluminescence sandwich immunoassay. GPNMB concentration was quantified in a solid phase sandwich ELISA using the R&D Systems Human Osteoactivin/GPNMB DuoSet ELISA kit. Methodological details are included in eMethods. Association between SOMAmer intensities on the one hand, and immunoassay-based measures of TNNT2 and GPNMB concentrations on the other, were assessed by Pearson correlation.

## RESULTS

### Participant characteristics in discovery and replication cohorts

The discovery cohort included 161 participants with ALS and 165 healthy controls (henceforth “ALS” and “controls”, respectively). After exclusion of samples that did not meet data QC criteria, the analysis dataset for plasma samples comprised 157 ALS and 163 controls. Longitudinal plasma and matching phenotypic data were available from 132 ALS and from 108 controls (Table 1). The analysis dataset for CSF samples comprised 70 ALS and 51 controls; longitudinal CSF and matching phenotypic data were available from 30 ALS and 24 controls (eTable 1). The replication cohort comprised 83 ALS and 28 controls. After data QC, 80 ALS and 28 controls were included in the analysis dataset for plasma, and 82 ALS and 25 controls in the analysis dataset for CSF (Table 1). Paired longitudinal phenotypic data and plasma (or CSF) samples were available from 37 participants with ALS (eTable 1).

**Table 1.** Participant Characteristics (Plasma)

|  |  |  | Discovery Cohort |  | Replication Cohort |  |
| --- | --- | --- | --- | --- | --- | --- |
|  |  |  | ALS<sup>a</sup> | Controls | ALS<sup>b</sup> | Controls |
| **Study Cohort** |  |  |  |  |  |  |
| Number of participants | N |  | 157 | 163 | 80 | 28 |
| Baseline age, years | Median (Q1-Q3) |  | 61 (54-68) | 49 (37-58) | 58 (51-66) | 46 (40-56) |
| Sex, male | N (%) |  | 74 (47%) | 68 (42%) | 44 (55%) | 21 (75%) |
| Baseline ALSFRS-R | Median (Q1-Q3) |  | 38 (32-42) | NA | 36 (31-41) | NA |
| Months since onset | Median (Q1-Q3) |  | 16.2 (7.7-32.7) | NA | 27.6 (18.1-38.2) | NA |
| Baseline ΔFRS, point(s)/month | Median (Q1-Q3) | | 0.6 (0.3-1.1) | NA | 0.4 (0-2.8) | NA |
| **Longitudinal Subset** |  |  |  |  |  |  |
| Number of participants | N |  | 132 | 108 | 37 | 0 |
| # of visits per person | Median (range) |  | 4 (2-14) | 5 (2-9) | 3 (2-3) | 1 |

ALSFRS-R = ALS Functional Rating Scale, Revised; $\Delta$ FRS = deltaFRS, calculated as (48 - baseline ALSFRS-R) / (months from onset to baseline); Q1 = 1<sup>st</sup> quartile; Q3 = 3<sup>rd</sup> quartile.
<sup>a</sup> Includes 35 *SOD1*, 16 *C9orf72*, 5 other genes, and 101 with no pathogenic variant identified.
<sup>b</sup> Includes 3 *C9orf72*.

### Disease state biomarkers in plasma and CSF

A total of 329 plasma proteins (345 SOMAmers) were significantly differentially regulated between ALS and controls (Benjamini Hochberg adjusted p-value < 0.05). Among them, 132 proteins (138 SOMAmers) were upregulated, and 197 proteins (207 SOMAmers) were downregulated, in ALS. Additionally, 4 of the differentially regulated proteins (5 SOMAmers) were more than 40% lower in ALS compared to controls (log2 fold-change [log2FC] < −0.5). Proteins with the greatest reduction in ALS included ART3 and ANTXR2, among others (Figure 1A, 1B). On the other hand, levels of 21 proteins (21 SOMAmers) were more than 40% elevated compared to controls. Proteins with the greatest elevation in ALS included PDLIM3, CA3, MYL11, APOBEC2, TNNT2, TNNI2, MYOM2, ACTN2, KLHL41 and HSPB6, among others (Figure 1A and 1C). Results were essentially unchanged irrespective of whether Plate ID was included as a fixed effect, a random effect or dropped entirely from the models. While the concentration of some proteins varied by sex, disease-related differential regulation of these proteins was not impacted by sex or underlying genotype (data not shown).

**Figure 1.**
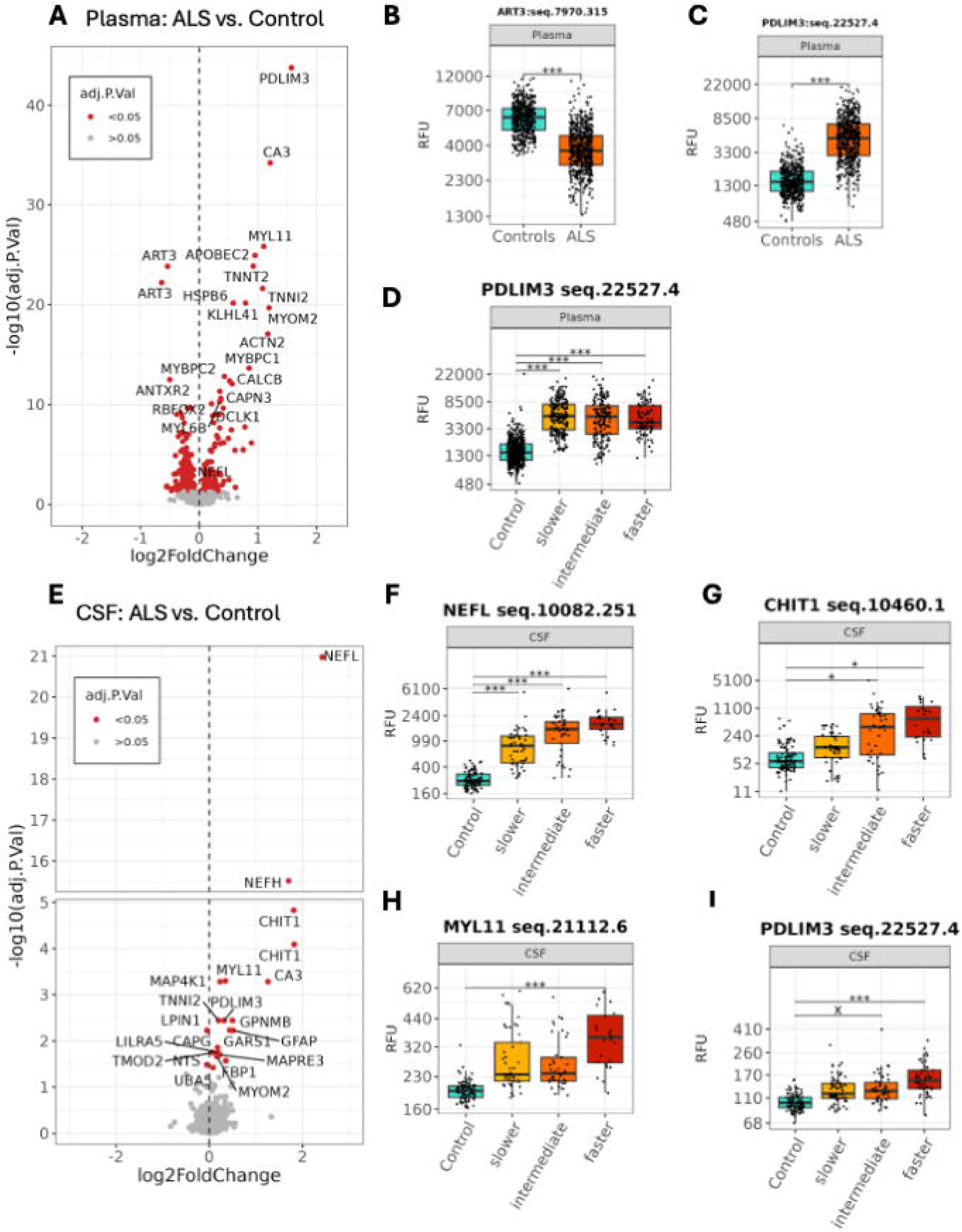
Plasma and CSF markers differentially regulated in ALS vs. healthy controls. (**A-D**) Plasma markers. (**A**) Volcano plot of ALS compared to healthy controls. SOMAmers in red have intensities that are statistically different. (**B-C**) Boxplots showing normalized age-, sex- and plate-adjusted SOMAmer intensities across controls and ALS for ART3 and PDLIM3. (**D**) Boxplot showing normalized age-, sex-, and plate-adjusted SOMAmer intensities for PDLIM3, in controls and ALS subgroups based on estimated disease progression rate. (**E-I**) CSF markers. (**E**) Volcano plot of ALS compared to healthy controls. SOMAmers in red have intensities that are statistically different. (**F-I**). Boxplots showing normalized age-, sex-, and plate-adjusted SOMAmer intensities for NEFL, CHIT1, MYL11 and PDLIM3, in controls and in ALS subgroups based on estimated progression rate. Adjusted P-value <0.001 denoted by ***; 0.001-0.01 denoted by **; 0.01-0.05 denoted by *; and 0.05-0.1 by “x”. RFU = relative fluorescence units. Seq.number, annotations of SOMAmers

A total of 20 CSF proteins (21 SOMAmers) were significantly differentially regulated between ALS and controls (Benjamini Hochberg adjusted p-value < 0.05). Most of these proteins were upregulated, with NEFL, NEFH, CHIT1, and CA3 being the most striking. MYL11, PDLIM3, GPNMB, and GFAP, among others, were more modestly upregulated (Figure 1E-I). Disease-associated regulation was not affected by sex or genotype (data not shown).

### Disease progression biomarkers in plasma

Plasma markers of disease progression were identified primarily based on their association with a longitudinal change in ALSFRS-R, a measure of functional impairment (Figure 2). Proteins that increase with disease progression, selected based on highest log2FC and smallest adjusted p-value, included TNNT2, PTN, PDLIM3, and MYL11. Conversely, proteins that decreased with disease progression, selected using the same criteria, included ANTXR2, CLEC3B, CRTAC1, ART3, and GDF11/MSTN (which share a high degree of amino acid sequence identity) (Figure 2A). Importantly, these candidate biomarkers remain consistent regardless of statistical method used (eFigure 3). For GDF11/MSTN, based on SOMAmers that recognize each protein individually, this signal appears to be driven by the muscle mass regulator myostatin (MSTN) (eFigure 4).

**Figure 2.**
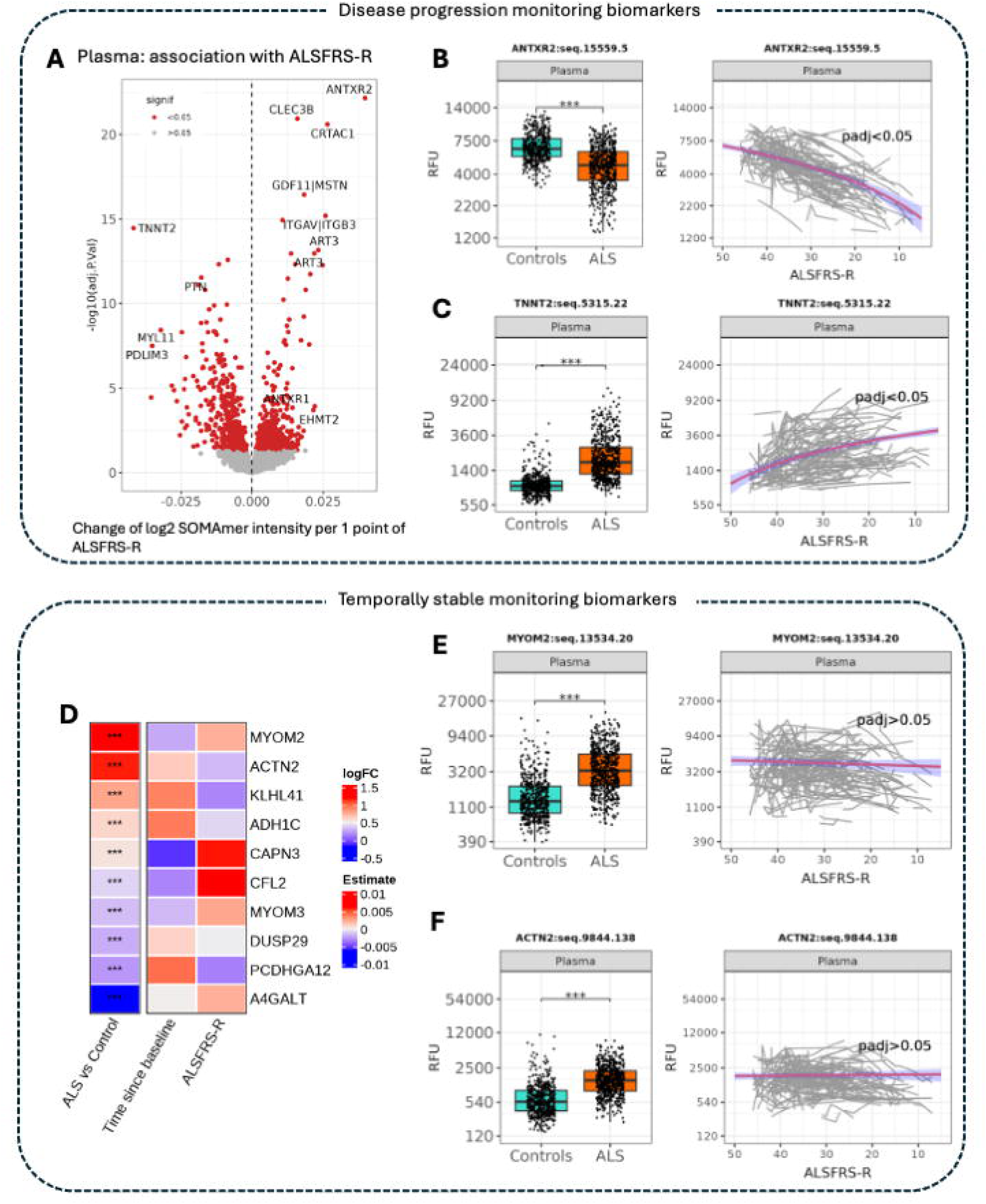
Monitoring biomarkers in plasma (A-C) Disease progression biomarkers. (**A**) Volcano plot showing adjusted p-values vs. change of log2 SOMAmer intensity per 1-point change in ALSFRS-R, estimated by LME. Negative values signify negative correlation, with protein (e.g. TNNT2, PTN, PDLIM3) levels increasing as ALSFRS-R declines (i.e. disease progresses). Positive values signify positive correlation, with protein (e.g. ANTXR2, CLEC3B, CRTAC1) levels decreasing as ALSFRS-R declines. SOMAmers marked in red are significantly associated with ALSFRS-R, with an adjusted p-value of <0.05. (**B-C**) Boxplots (comparing ALS to controls) and spaghetti plots (SOMAmer intensity vs. ALSFRS-R) of ANTXR2 and TNNT2, as two illustrative examples. **(D-F) Temporally stable biomarkers**. (**D**) Heatmap showing regression coefficients for top 10 examples of proteins, whose matching SOMAmers meet the dual criteria of having the greatest log-fold change in SOMAmer intensity in ALS compared to controls AND remaining relatively stable (i.e. no evidence for significant change) over time and across disease progression. The left panel shows log2FoldChange indicating difference of mean protein level in plasma of ALS vs control, while the right panel shows regression coefficients (Estimate) for change of protein level on log2 scale per 1 month disease duration since baseline and the same change per 1 unit of ALSFRS-R (2^nd^ and 3^rd^ columns, respectively). (**E-F**) Boxplots (comparing ALS to controls) and spaghetti plots (SOMAmer intensity vs. ALSFRS-R) of MYOM2 and ACTN2, as two illustrative examples. RFU = relative fluorescence units. padj = adjusted p-values.

**Figure 3.**
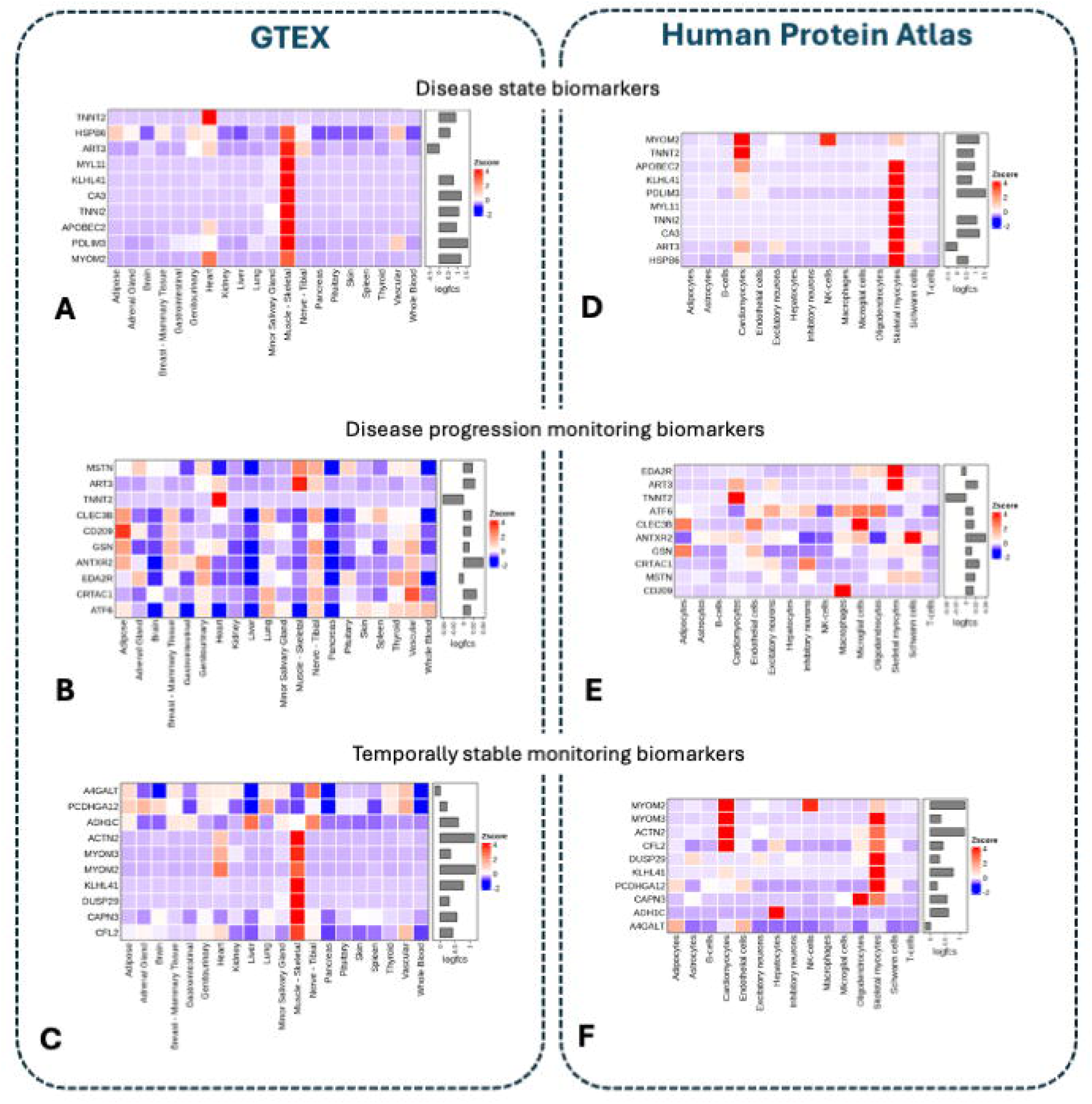
Plasma biomarkers are encoded by muscle-enriched genes. Left and right panels show results of GTEX- and Human Protein Atlas (HPA)-based enrichment analysis. (**A-C**) Heatmaps showing scaled mean transcript per million (TPM) values for each organ for genes encoded by plasma biomarkers selected by lowest adjusted p-value. Sidebars on the right indicate effect size based on log2FC in SOMAmer intensity, comparing ALS vs. controls (**A**, **C**) and per unit change in ALSFRS-R (**B**). (**D-F**) Heatmaps showing scaled normalized TPM values for plasma biomarkers selected by lowest adjusted p-value in the selected cell types. Sidebars on the right indicate effect size based on log2FC in SOMAmer intensity, comparing ALS vs. controls (**D**, **F**) and per unit change in ALSFRS-R (**E**). The log2FC value for MSTN corresponds to MSTN|GDF11 SOMAmer seq.2765.4 (**E**).

**Figure 4.**
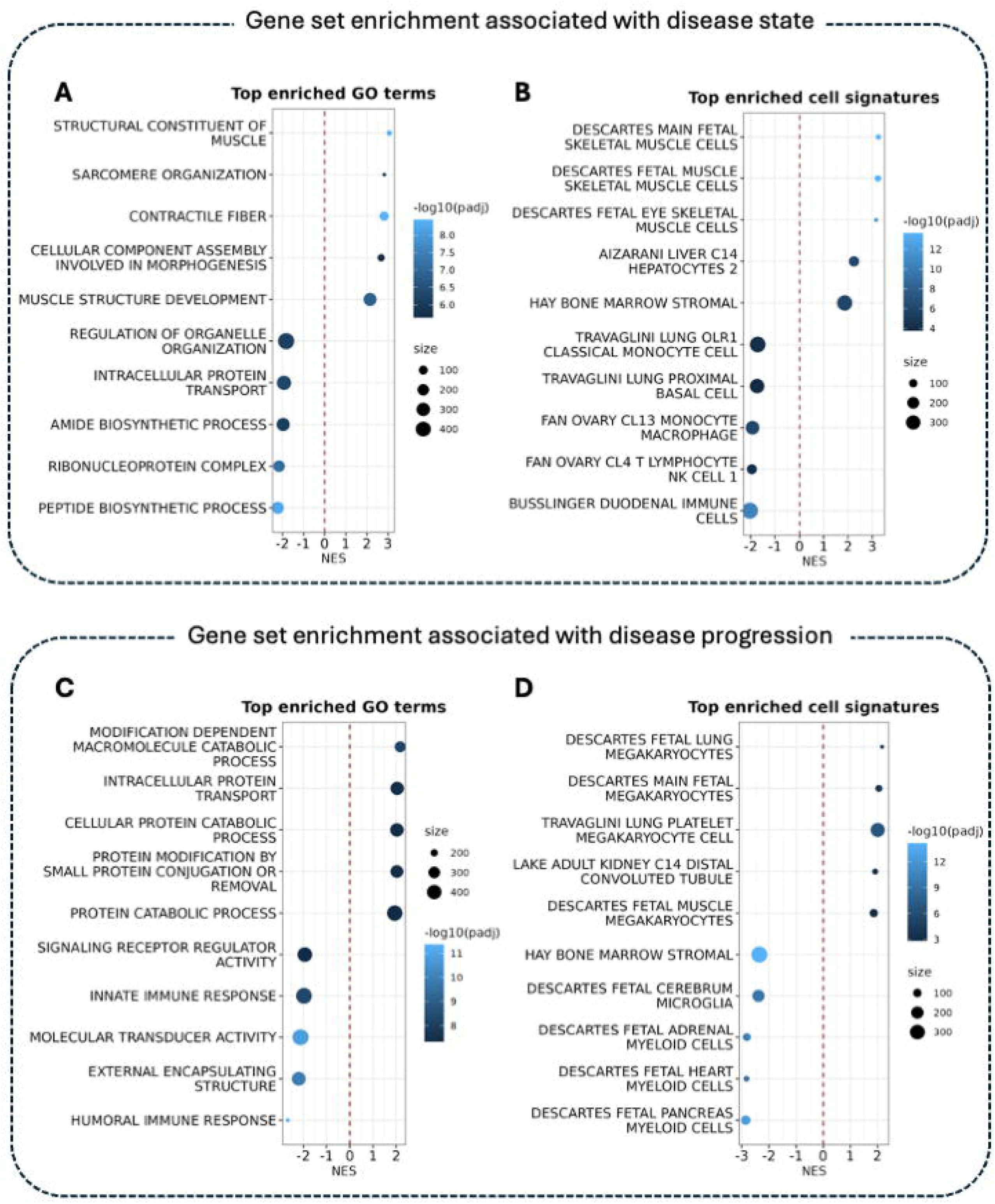
Gene ontology-based enrichment. Skeletal muscle signature in plasma is among the top enriched gene sets differentiating ALS vs. controls, while immune/inflammatory features are among the top signatures associated with disease progression. Enrichment plots show top 5 upregulated and top 5 downregulated Gene Ontology (GO) terms and cell type-specific signatures in plasma SOMAmers that are differentially regulated in ALS vs. controls (**A**, **B**), and associated with ALSFRS-R (**C**, **D**). NES = normalized enrichment score. Dot size represents the number of proteins overlapping with a gene signature. Note that the sign for NES in **A** and **B** coincides with the directional change of protein level in ALS vs. controls, with a positive NES indicating upregulation of a given gene signature, and a negative NES indicating depression of a given gene signature. By contrast, the sign for NES in **C** and **D** coincides with the ALSFRS-R, with a negative NES reflecting genes associated with a lower ALSFRS-R (i.e. more advanced disease).

Similar plasma markers were consistently identified (e.g. TNNT2, PTN, PDLIM3, MYL11, ANTXR2, ART3, CLEC3B and GDF11/MSTN) irrespective of association with ALSFRS-R or follow-up duration (Figure 2A, eFigure 5). The trajectories of the candidate markers of disease progression are illustrated for ANTXR2 (Figure 2B) and TNNT2 (Figure 2C). To illustrate the potential utility of these biomarkers of disease progression, we indexed their rates of change to the standard error (namely, the t-value from the mixed effect models) and compare them to similar estimate for ALSFRS-R. On average ALSFRS-R declines by −0.72 points/month (SE 0.05) with a t-value of −13.7. For ANTXR2 and TNNT2, rates of change are 11.2 (0.49) and −7.3 (0.48) units (log2 scale)/month, respectively, with t-values of 22.6 and −15.1.

### Temporally stable biomarkers in plasma

Markers that were elevated in ALS compared to controls but remained relatively stable were not associated with time since baseline or with longitudinal change in ALSFRS-R (Figure 2D). Top candidates for such markers included MYOM2 (Figure 2E), ACTN2 (Figure 2F), and KLHL41. There was, however, substantial within-person variability in the relative concentration of these markers, which might limit their utility as potential response markers.

### Prognostic biomarkers in plasma and CSF

In an unbiased analysis evaluating the relationship between baseline SOMAmer intensity and the subsequent rate of disease progression as measured by ALSFRS-R, no novel prognostic biomarker candidates were identified in plasma or CSF. When comparing proteins in patients with faster versus slower disease progression, however, some plasma proteins were modestly upregulated (e.g., PLIN3) while others were modestly downregulated (e.g., CLEC3B, CRTAC1); but these did not remain significant after p-value adjustment for multiple testing (data not shown). As expected, in CSF, NEFL SOMAmer intensity was greatest in those with faster disease progression compared to those with slower disease progression, although statistically significant differences were only apparent when compared to controls (Figure 1F). The same was true for CSF CHIT1 and MYL11 (Figure 1G, 1H), and with similar trends for PDLIM3 (Figure 1I), GPNMB, GFAP and CA3 (eFigure 6).

### Replication in an independent cohort

Candidate plasma and CSF biomarkers identified in the discovery cohort were largely replicated in an independent, albeit smaller, cohort (eFigure 7). In the replication cohort, lead plasma candidates identified as elevated in ALS compared to controls included PDLIM3, CA3, MYOM2, MYL11, TNNT2, and APOBEC2, among others. Lead plasma candidates identified as lower in ALS included ART3 and ANTXR2. Lead biomarker candidates that were temporally stable in plasma included MYOM2, MYOM3 and CAPN3, whereas lead plasma biomarker candidates of disease progression included TNNT2, PDLIM3 and MYL11 (increasing), as well as ART3, ANTXR2 and CRTAC1 (decreasing). In CSF, CHIT1, NEFL, NEFH, and GPNMB were replicated as elevated in ALS vs. controls. A few biomarker candidates in plasma (e.g. EDA2R) and CSF (e.g. CA3) were not consistent across the discovery and replication cohorts.

### Biomarker confirmation through targeted immunoassay

We observed a highly significant correlation between plasma SOMAmer intensity and TNNT2 concentration (r=0.81, p<0.0001, eFigure 8B). In the ALS group, median (25^th^-75^th^ percentile) plasma TNNT2 concentration at baseline was 19ng/L (12-37) compared to 7ng/L (5-9) among controls (p<0.0001, eFigure 8B), and TNNT2 concentration increased by an average (95% CI) of 0.44ng/L per month (0.24-0.64) (eFigure 8C). Similarly, we observed a highly significant correlation between plasma SOMAmer intensity and GPNMB concentration (r=0.66-0.93, depending on individual SOMAmer, with p<0.0001, eFigure 9A). In the ALS group, median (25^th^ – 75^th^ percentile) plasma GPNMB concentration at baseline was 8.4ng/mL (6.9-10.9) compared to 4.4ng/mL (1.7-7.3) among controls (p=0.011, eFigure 9B). On average, GPNMB concentrations were relatively stable over time (eFigure 9C).

### Biological implications

In a Genotype-Tissue Expression (GTEx)-based analysis, plasma biomarkers with adjusted p-values <0.05 were linked to their putative organ of origin based on mRNA expression levels. An unbiased analysis plotting average log2FC vs. tau (a tissue specificity index), revealed that proteins with the largest average log2FC and highest specificity were encoded by skeletal muscle-specific genes. This observation held true for disease state (i.e. ALS vs. control) and temporally stable biomarkers, but not for disease progression biomarkers (eFigure 10). The enrichment in skeletal muscles was further illustrated based on the organ distribution of the top hits for disease state and temporally stable biomarkers (Figure 3A, 3C); and more diverse tissue origins among top hit biomarkers of disease progression (Figure 3B).

At a cellular level (based on the Human Proteome Atlas [HPA]), the top 10 proteins with highest statistically significant relative fold-change in ALS vs. controls are expressed almost exclusively in skeletal myocytes (Figure 3D). The same was true for candidate temporally stable biomarkers, with contribution from oligodendrocytes (CAPN3), hepatocytes (ADH1C) and Natural Killer cells (MYOM2) (Figure 3F). Biomarkers of disease progression were highly expressed in cardiac myocytes (TNNT2), skeletal muscle (ART3), microglia (CLEC3B), macrophages (CD209), and Schwann cells (ANTXR2) (Figure 3E). Importantly, these enrichment analyses were based on the expression atlas of healthy tissues and might differ for some genes in the context of diseased tissue or in response to environmental factors.

The enrichment analyses based on molecular function, cellular component, and biological processes (Gene Ontology) as well as cell type signature (Human Molecular Signatures Database) similarly implicate skeletal muscle. Gene set enrichment analysis confirmed that muscle pathology is the dominant biological theme (Figure 4A-B). These analyses also revealed an increased immune/inflammatory signal in association with disease progression (Figure 4C-D).

## DISCUSSION

This ALS longitudinal proteomic study is unique, to date, insofar as it includes a large discovery cohort as well as a moderately sized replication cohort. In contrast, prior studies have been limited to cross-sectional samples from much smaller cohorts, in the range of 10-42 ALS and 5-30 controls.^22–26^ Unlike most prior proteomic studies that focused exclusively on differentiating ALS vs. controls, our analysis strategy included consideration of prognostic and monitoring (both disease progression and temporally stable) contexts of use.

Among the ∼7,000 SOMAmers measured, upregulated plasma markers in ALS versus controls most often reflect skeletal muscle biology, based on pathway analysis and tissue specificity. These proteins are also candidate biomarkers of disease progression: TNNT2, PDLIM3, and MYL11 increase significantly with disease progression, while ANTXR2, ART3, CLEC3B, and MSTN decrease significantly as the disease unfolds. Importantly, these findings were replicated in an independent cohort, and one of these markers (TNNT2) was analytically confirmed using the Roche immunoassay, providing strong evidence for its potential clinical utility. Skeletal muscle tissue has also been identified as a potential source of biomarkers, based on clinicopathological correlation with disease progression and evidence of similar dysfunction in the *SOD1*^G93A^ ALS mouse model.^27,28^ Moreover, while largely categorized as of skeletal muscle origin, some of the monitoring biomarkers we identified (e.g. ANTXR2 and PDLIM3) are either also expressed in, or interact with, extracellular matrix (ECM) components (eTable 2), and have been described in molecular pathways linked to inflammation, as well as tissue ageing and cell senescence.^29^

In CSF, we confirmed prior observations that NEFL, NEFH and CHIT1 are elevated in ALS.^10,11,13,30,31^ The differential abundance of neurofilament proteins (e.g., NEFL) observed in CSF was less prominent in plasma, consistent with our other SomaScan profiling studies (data not shown). This may be attributable to non-specific binding of the SOMAmer or potential interference within the plasma matrix. In addition, we identified CA3 and MYL11, the expression of which is enriched in skeletal muscle; as well as GPNMB, which is of multi-cellular origin and involved in synthesis of ECM components, and which exerts neuroprotective anti-inflammatory effects through a broad spectrum of immune cells including astrocytes.^32^

Until now, there have been limited biofluid markers for ALS disease progression, which might be used as outcome measures in early phase clinical trials. Our approach to biomarker discovery has identified and replicated an array of novel candidates that change over time as the disease progresses. Based on the rates of change of these biomarkers relative to their standard errors, they compare favorably to the ALSFRS-R, offering the added value of reliable quantification with less potential for evaluator bias. Each of these candidates will require further analytical confirmation using suitable orthogonal bioanalytical methods, such as immunoassays. Our identification of TNNT2 as a plasma marker that increases over time during the clinically manifest stage of disease is consistent with prior observations,^28,33–36^ including a small SomaScan study.^23^ TNNT2 is a structural protein integral to contractile properties of striated muscle. While TNNT2 is primarily expressed in cardiac muscle (both embryonic and adult), it is also transiently expressed in skeletal muscle during development^37^ and following denervation.^38^ Moreover, it has been reported that TNNT2 is re-expressed in diseased skeletal muscle in patients with a wide range of neuromuscular disorders.^39–41^ These observations are consistent with the known biology of skeletal muscle in which regeneration, a recognized source of TNNT2, is a response to injury. Indeed, muscle tissue expression of TNNT2 is increased in the *SOD1* mouse model of ALS^42^ and in people with ALS.^28^ Notwithstanding an incomplete understanding of the biological mechanism, our data suggest that plasma TNNT2 could be considered in pharmacological clinical trials to monitor the possible effects of therapeutic interventions. Interestingly, the decrease in MSTN (myostatin, a negative regulator of muscle growth) suggests a compensatory response, consistent with skeletal muscle transcriptional down regulation of MSTN.^28^

The increase in PDLIM3 in ALS is of particular interest given the possibility that this might be related to loss of TDP-43 nuclear function, with resulting aberrant splicing.^43^ Single cell RNA-seq data of post-mortem FTD-ALS human brain showed that loss of nuclear TDP-43 is associated with widespread transcriptional changes, including upregulation of PDLIM3.^44^ Moreover, muscle biopsies from autosomal dominant rimmed vacuole myopathy patients with a frameshift mutation in *TARDBP* (the gene encoding the TDP-43 protein) displayed altered splicing patterns of pre-mRNAs encoding sarcomeric proteins (including PDLIM3) and upregulation of muscle regeneration genes.^45^ The basis for changes in proteins such as ANTXR2 (Schwann cells), CAPN3 (oligodendrocytes), CD209 (macrophages), and MYOM2 (NK cells) is unclear, but might reflect an increasing immune/inflammatory cell signal in association with disease progression.

The SomaScan high-throughput proteomics assay employed in this study provided us with a unique opportunity to deeply profile the ALS disease proteome longitudinally and to identify novel biomarker candidates that may have remained undiscovered through other methods. While the aptamer-based proteomics proved to be a powerful tool for novel biomarker discovery at scale with the ability to measure thousands of proteins simultaneously, it is not without its limitations. The technology relies on aptamer-based assays, which provide only relative expression levels. Additionally, the potential for alterations to protein structure through oligomerization, degradation, post-translational modifications, or genetic polymorphisms could significantly and unpredictably alter binding affinity and quantification. Therefore, clinical application of these biomarkers will require more targeted technologies to ensure specificity and to permit absolute quantification. We already confirmed one of the top muscle marker candidates, TNNT2, and one of our top CSF hits, GPNMB. Similar efforts are underway for additional biomarker confirmation, which will further advance our candidates towards a clinical application.

Strengths of this study include its use of a large discovery cohort with meticulously collected phenotypic data, as well as a replication cohort, albeit smaller in size. Also, our approach to biomarker discovery deviated from the more traditional approach of simply comparing ALS to controls. Instead, our analysis was predicated upon an understanding of the potential biomarker context of use. For example, to identify biomarkers of disease progression, we prioritized those with the strongest association with longitudinal measures of ALSFRS-R. Similarly, we sought to identify markers that are elevated or depressed in ALS relative to controls, but with relative stability over time, akin to monitoring biomarkers such as NfL. This approach has yielded a host of new biomarker candidates and underscores the importance of skeletal muscle biology. It has also implicated other biological mechanism related to inflammation and the potential role of the ECM in the unraveling of innate and adaptive immunological responses as characteristic features of disease. Importantly, our analytic approach to biomarker discovery has revealed the likely context of use for the biomarker candidates identified.

## Supporting information

eMethods

eTable 1

eTable 2

eTable 3

eTable 4

eFigure 1

eFigure 2

eFigure 3

eFigure 4

eFigure 5

eFigure 6

eFigure 7

eFigure 8

eFigure 9

eFigure 10

## Acknowledgements

The authors thank participants in the CReATe Consortium’s Phenotype-Genotype-Biomarker (PGB1) study and TRIAL READY study at the University of Miami site; participants in the Clinical Research in ALS (CRiALS) study at the University of Miami; the research team at the University of Miami; the CReATe Consortium and CRiALS project management and data management teams, genomic cores, and biorepositories; the Rare Diseases Clinical Research Network Data Management and Coordinating Center (RDCRN DMCC); the NEALS biorepository, including the Operational Director, Becky Fillingham, and the NEALS Consortium Operational Leadership, participants who contributed samples to the NEALS biorepository; and Kezia Allen and Eliza Lin for their assistance with TNNT2 immunoassays. We also thank the donors of the Novartis Basel Tissue Donor Program and the DARe department samples management team for technical support. Lastly, we thank the contracting teams from each institution represented by our co-authors for their support in facilitating this collaborative work.

## Data availability

Request should be sent to the corresponding author. The full SomaScan Discovery and Replication datasets are available upon reasonable request via a Data Transfer Agreement (DTA) with Novartis. Associated phenotypic data for the Discovery and Replication cohorts are available upon reasonable request to, respectively, the University of Miami and the Massachusetts General Hospital (on behalf of NEALS), via DTA.

## Supplementary material

Supplementary material is available online.

## REFERENCES

1. Bensimon G, Lacomblez L, Meninger V, and the ALS/Riluzole study group. A controlled trial of riluzole in amyotrophic lateral sclerosis. New England Journal of Medicine 1994; 330: 585–91.

2. The Writing Group on behalf of the Edaravone (MCI-186) ALS 19 Study Group. Safety and efficacy of edaravone in well defined patients with amyotrophic lateral sclerosis: a randomised, double-blind, placebo-controlled trial. Lancet Neurol 2017; 16(7): 505–12.

3. Miller TM, Cudkowicz ME, Genge A, et al. Trial of Antisense Oligonucleotide Tofersen for SOD1 ALS. The New England journal of medicine 2022; 387(12): 1099–110.

4. Benatar M, Boylan K, Jeromin A, et al. ALS biomarkers for therapy development: State of the field and future directions. Muscle & nerve 2016; 53(2): 169–82.

5. FDA-NIH Biomarker Working Group. BEST (Biomarkers, EndpointS, and other Tools) Resource. Silver Spring (MD): Food and Drug Administration (US). Co-published by National Institutes of Health (US), Bethesda (MD); 2016.

6. Shepheard S, Wuu J, Cardoso M, et al. Urinary p75 extracellular domain; A biomarker for prognosis, progression and pharmacodynamic effect in ALS. Neurology 2017; 88(12): 1137–43.

7. Winer L, Srinivasan D, Chun S, et al. SOD1 in cerebral spinal fluid as a pharmacodynamic marker for antisense oligonucleotide therapy. JAMA neurology 2013; 70(2): 201–7.

8. Gendron TF, Chew J, Stankowski JN, et al. Poly(GP) proteins as pharmacodynamic biomarker for C9ORF72 associated amyotrophic lateral sclerosis. Science Translational Medicine 2017; 9: 1–12.

9. Lesterhuis WJ, Bosco A, Millward MJ, Small M, Nowak AK, Lake RA. Dynamic versus static biomarkers in cancer immune checkpoint blockade: unravelling complexity. Nat Rev Drug Discov 2017; 16(4): 264–72.

10. Benatar M, Zhang L, Wang L, et al. Validation of serum neurofilaments as prognostic and potential pharmacodynamic biomarkers for ALS. Neurology 2020; 95(1): e59–e69.

11. Thompson AG, Gray E, Verber N, et al. Multicentre appraisal of amyotrophic lateral sclerosis biofluid biomarkers shows primacy of blood neurofilament light chain. Brain Commun 2022; 4(1): fcac029.

12. Benatar M, Macklin EA, Malaspina A, et al. Prognostic clinical and biological markers for amyotrophic lateral sclerosis disease progression: validation and implications for clinical trial design and analysis. EBioMedicine 2024; 108: 105323.

13. Benatar M, Wuu J, Turner MR. Neurofilament light chain in drug development for amyotrophic lateral sclerosis: a critical appraisal. Brain : a journal of neurology 2023; 146(7): 2711–6.

14. Tran H, Moazami MP, Yang H, et al. Suppression of mutant C9orf72 expression by a potent mixed backbone antisense oligonucleotide. Nature medicine 2022; 28(1): 117–24.

15. Fader KA, Pardo ID, Kovi RC, et al. Circulating neurofilament light chain as a promising biomarker of AAV-induced dorsal root ganglia toxicity in nonclinical toxicology species. Mol Ther Methods Clin Dev 2022; 25: 264–77.

16. van den Berg LH, Rothstein JD, Shaw PJ, et al. Safety, tolerability, and pharmacokinetics of antisense oligonucleotide BIIB078 in adults with C9orf72-associated amyotrophic lateral sclerosis: a phase 1, randomised, double blinded, placebo-controlled, multiple ascending dose study. Lancet Neurol 2024.

17. Gentile JE, Heiss C, Corridon TL, et al. Evidence that minocycline treatment confounds the interpretation of neurofilament as a biomarker. medRxiv 2024.

18. Benatar M, McDermott C, Turner MR, van Eijk RPA. Rethinking phase 2 trials in amyotrophic lateral sclerosis. Brain : a journal of neurology 2025; 148(4): 1106–11.

19. Candia J, Daya GN, Tanaka T, Ferrucci L, Walker KA. Assessment of variability in the plasma 7k SomaScan proteomics assay. Sci Rep 2022; 12(1): 17147.

20. Emilsson V, Ilkov M, Lamb JR, et al. Co-regulatory networks of human serum proteins link genetics to disease. Science 2018; 361(6404): 769–73.

21. Kaiser S, Zhang L, Mollenhauer B, et al. A proteogenomic view of Parkinson’s disease causality and heterogeneity. NPJ Parkinsons Dis 2023; 9(1): 24.

22. Bereman MS, Beri J, Enders JR, Nash T. Machine Learning Reveals Protein Signatures in CSF and Plasma Fluids of Clinical Value for ALS. Sci Rep 2018; 8(1): 16334.

23. Berrone E, Chiorino G, Guana F, et al. SOMAscan Proteomics Identifies Novel Plasma Proteins in Amyotrophic Lateral Sclerosis Patients. International journal of molecular sciences 2023; 24(3).

24. He L, Zhou Q, Xiu C, et al. Circulating proteomic biomarkers for diagnosing sporadic amyotrophic lateral sclerosis: a cross-sectional study. Neural Regen Res 2024; 19(8): 1842–8.

25. Katzeff JS, Bright F, Lo K, et al. Altered serum protein levels in frontotemporal dementia and amyotrophic lateral sclerosis indicate calcium and immunity dysregulation. Sci Rep 2020; 10(1): 13741.

26. Xu Z, Lee A, Nouwens A, Henderson RD, McCombe PA. Mass spectrometry analysis of plasma from amyotrophic lateral sclerosis and control subjects. Amyotroph Lateral Scler Frontotemporal Degener 2018; 19(5-6): 362–76.

27. King PH. Skeletal muscle as a molecular and cellular biomarker of disease progression in amyotrophic lateral sclerosis: a narrative review. Neural Regen Res 2024; 19(4): 747–53.

28. Tsitsipatis D, Mazan-Mamczarz K, Si Y, et al. Transcriptomic analysis of human ALS skeletal muscle reveals a disease-specific pattern of dysregulated circRNAs. Aging (Albany NY) 2022; 14(24): 9832–59.

29. Zhang W, Liu Y, Zhang H. Extracellular matrix: an important regulator of cell functions and skeletal muscle development. Cell Biosci 2021; 11(1): 65.

30. Steinacker P, Verde F, Fang L, et al. Chitotriosidase (CHIT1) is increased in microglia and macrophages in spinal cord of amyotrophic lateral sclerosis and cerebrospinal fluid levels correlate with disease severity and progression. J Neurol Neurosurg Psychiatry 2018; 89(3): 239–47.

31. Thompson AG, Gray E, Bampton A, Raciborska D, Talbot K, Turner MR. CSF chitinase proteins in amyotrophic lateral sclerosis. J Neurol Neurosurg Psychiatry 2019; 90(11): 1215–20.

32. Neal ML, Boyle AM, Budge KM, Safadi FF, Richardson JR. The glycoprotein GPNMB attenuates astrocyte inflammatory responses through the CD44 receptor. J Neuroinflammation 2018; 15(1): 73.

33. Casmiro M, Graziani A. Serum troponin T in patients with amyotrophic lateral sclerosis. Acta Neurol Belg 2019; 119(2): 285–8.

34. Mach L, Konecny T, Helanova K, et al. Elevation of cardiac troponin T in patients with amyotrophic lateral sclerosis. Acta Neurol Belg 2016; 116(4): 557–64.

35. Castro-Gomez S, Radermacher B, Tacik P, Mirandola SR, Heneka MT, Weydt P. Teaching an old dog new tricks: serum troponin T as a biomarker in amyotrophic lateral sclerosis. Brain Commun 2021; 3(4): fcab274.

36. Klappe U, Chamoun S, Shen Q, et al. Cardiac troponin T is elevated and increases longitudinally in ALS patients. Amyotroph Lateral Scler Frontotemporal Degener 2022; 23(1-2): 58–65.

37. Anderson PA, Greig A, Mark TM, et al. Molecular basis of human cardiac troponin T isoforms expressed in the developing, adult, and failing heart. Circ Res 1995; 76(4): 681–6.

38. Rasmussen M, Jin JP. Troponin Variants as Markers of Skeletal Muscle Health and Diseases. Front Physiol 2021; 12: 747214.

39. du Fay de Lavallaz J, Prepoudis A, Wendebourg MJ, et al. Skeletal Muscle Disorders: A Noncardiac Source of Cardiac Troponin T. Circulation 2022; 145(24): 1764–79.

40. Rittoo D, Jones A, Lecky B, Neithercut D. Elevation of cardiac troponin T, but not cardiac troponin I, in patients with neuromuscular diseases: implications for the diagnosis of myocardial infarction. J Am Coll Cardiol 2014; 63(22): 2411–20.

41. Schmid J, Liesinger L, Birner-Gruenberger R, et al. Elevated Cardiac Troponin T in Patients With Skeletal Myopathies. J Am Coll Cardiol 2018; 71(14): 1540–9.

42. Saris CG, Groen EJ, van Vught PW, et al. Gene expression profile of SOD1-G93A mouse spinal cord, blood and muscle. Amyotroph Lateral Scler Frontotemporal Degener 2013; 14(3): 190–8.

43. Donde A, Sun M, Ling JP, et al. Splicing repression is a major function of TDP-43 in motor neurons. Acta neuropathologica 2019; 138(5): 813–26.

44. Liu EY, Russ J, Cali CP, Phan JM, Amlie-Wolf A, Lee EB. Loss of Nuclear TDP-43 Is Associated with Decondensation of LINE Retrotransposons. Cell reports 2019; 27(5): 1409–21 e6.

45. Ervilha Pereira P, Schuermans N, Meylemans A, et al. C-terminal frameshift variant of TDP-43 with pronounced aggregation-propensity causes rimmed vacuole myopathy but not ALS/FTD. Acta neuropathologica 2023; 145(6): 793–814.

