## Supplementary material for "Skeletal muscle biomarkers of amyotrophic lateral sclerosis: a large-scale, multi-cohort proteomic study": eMethods

### Initial data processing

Initial quality assessment and basic raw SomaScan® data processing was performed using a SomaLogic developed analysis pipeline. After extracting the array intensity, control probes were removed from the dataset. For CSF and plasma samples, raw intensities were normalized using adaptive normalization by maximum likelihood (ANML) (SomaLogic). Intensity values were log2-transformed. After standard data processing, 7288 SOMAmers matching 6401 proteins (by Uniprot ID) corresponding to 6400 human genes analytes in plasma and CSF remained for analysis. All 7288 SOMAmers were retained for downstream statistical analysis. Samples with SOMAmer intensity distributions flagged as outlier were excluded from downstream analysis [1] .

### Exploratory analysis

An exploratory analysis was undertaken prior to formal statistical analysis. For each matrix (plasma or CSF), Principal Component Analysis (PCA) was performed on all available samples including buffer and calibrator samples. Plasma and CSF samples were analyzed separately. No major batch effects were identified by PCA. Outlier samples detected by PCA were labeled as outliers and excluded by AutoQC pipeline (eFigure 2). To further characterize the variation in the proteome data, PCA was re-run *after* calibrator and buffer samples removal as part of QC. Several biological and technical co-variates driving variance were identified for both plasma and CSF. To assess the associations between principal components and these biological (age at collection, sex, disease status [ALS vs. control]) and technical (plate) co-variates, we employed simple linear regression whereby each of the first 10 principal components was modeled as the dependent variable and each co-variate as independent variables (e.g., PC1 ~ age at collection or PC2 ~ sex). Outputs of all tests were combined, and raw p-values were adjusted with the Bonferroni method to account for multiple testing. The association between a principal component and a co-variate was considered significant if the Bonferroni-adjusted $p$-value $< 0.05$. Several biological and technical co-variates driving variance were identified in both plasma and CSF datasets including sex, age at sample collection, and disease state. We also identified SOMAscan hemoglobin levels as a source of variance in our study, indicating hemolysis in some plasma samples or blood-contamination of some CSF samples. Correlations between CSF and plasma proteins with hemoglobin levels are summarized in eTable 3.

**Patient categorization based on rate of disease progression**

To stratify participants with ALS based on rates of disease progression, we used a multivariate LME modeling approach. Briefly, the following LME was fitted to the longitudinal data on 132 unique individuals with at least 2 distinct ALSFRS-R measurements to estimate the expected rate of ALSFRS-R decline per year for each person: ALSFRS-R ~ Sex*YrSinceOs_ALS + CollAge+GeneGrp*YrSinceOs_ALS + (YrSinceOs_ALS|UIDx), where YrSinceOs_ALS is years from ALS symptom onset to sample collection, CollAge is the age at time of sample collection, GeneGrp reflects the 4 genotype subgroups (*SOD1*, *C9orf72*, other pathogenic genes, and no identifiable pathogenic variant), and UIDx is the unique participant identifier. Based on predicted rates of ALSFRS-R decline, participants with ALS were categorized into faster (>1 point/month), intermediate (0.5-1 point/month), and slower progressors (<0.5 point/month).

**Group comparisons**

All log2-transformed SOMAmer intensities were compared between healthy controls and people with ALS as well as across progression subgroups (faster vs. intermediate vs. slower) using a linear mixed-effects (LME) model with a subject-specific random intercept to account for the intrasubject correlation structure of multiple measurements over time [2]. To fit linear mixed effect models lmerTest R package was employed [3]. All models were adjusted for age, sex, plate, and hemoglobin level. To conduct all possible pairwise comparisons between groups (faster vs slower, slower vs intermediate etc.), emmeans R package was used [2]. Benjamini–Hochberg procedure was used to adjust for comparison across multiple hypotheses.

To corroborate our findings based on LME modeling, we also employed empirical Bayes standard deviation shrinkage method, as implemented in the limma package [4] . We conducted analysis two ways, by including (a) all samples, with blocking by participant ID to account for non-independence of samples (correlated variance), and (b) only the first sample from each person. All limma models were sex, age, plate ID and hemoglobin levels adjusted. Benjamini–Hochberg procedure was used to adjust for comparison across multiple hypotheses.

**Modeling longitudinal change in protein levels**

Longitudinal trends for each individual SOMAmer were modeled as a constant rate of change (i.e. linear slope) over time. Specifically, an LME analysis was performed with analyte log2 concentration as the dependent variable, either follow-up duration or ALS Functional Rating Scale–Revised (ALSFRS-R) as the independent variable, and a subject-specific random intercept and slope to account for potential differences across individuals. An unstructured covariance matrix was assumed between the subject-specific random intercept and slope parameters. The outcomes of all statistical tests (summary statistics) from group comparisons and association of protein levels with continuous variables can be found in eTable 3.

**Temporally stable biomarkers**

Temporally stable biomarkers are those that are elevated or decreased in people with clinically manifest ALS compared to a healthy control population, but stable over time. These biomarker candidates were identified based on significant difference from healthy controls (adjusted p-value<0.05), but no evidence of longitudinal changes based on association with ALSFRS-R and years since symptom onset (adjusted p-values > 0.15).

**Enrichment analyses**

To further our understanding of the biological implications and to gain insights into the potential sources of the identified disease state biomarker candidates, we conducted a comprehensive organ (Genotype-Tissue Expression [GTEx]) and cell type (Human Proteome Atlas [HPA]) expression enrichment analyses, supplemented by functional process enrichment studies.

**Organ and cell type expression enrichment analysis**

GTEx v10 and Human Protein Atlas (HPA) version 23.0 and Ensembl version 109 were used to infer organ and cell type specificity of gene expression. Organ specificity was quantified using the tau parameter [5], where organ or tissue specific genes have tau>0.85, and genes expressed in multiple organs have tau<0.85 [5].

**Gene set enrichment analysis**

Gene set enrichment analysis [6] was conducted with fgsea R package [7]. SOMAmers were pre-ranked based on t-statistics estimated by LME. In the event of multiple SOMAmers mapped to a single entrez gene ID, maximum absolute T-statistic values were used for ranking. Gene sets v2024.1.Hs was downloaded from the Broad institution [www.gsea-msigdb.org]. The top 10 enriched signatures (5 upregulated and 5 downregulated) from Gene Ontology (C5) and cell type signature (C8) gene set were plotted with custom R script (eTable4).

**Data visualization**

All data visualizations were generated using the ggplot2 package (Wickham, 2016) unless stated otherwise. Boxplots and spaghetti plots show age-, sex-, plate-adjusted ANML-normalized values. Briefly, linear associations of each SOMAmer ANML-normalized intensities to age at collection, sex and plate were regressed out with RemoveBatchEffect function from limma package to obtain values free of the potentially confounding effect of those 3 co-variates. Adjusted p-values (BH-corrected) obtained from lmerTest were converted using the stars.pval function from the gtools R package. All figures are based on the output of mixed effect models. The limma output was used only as a confirmation for single-timepoint analysis, and can be found in eTable 1.

**TNNT2 Immunoassay validation**

TNNT2 (Troponin T2) concentration was quantified using the Roche Elecsys Troponin T high sensitivity (hs) electrochemiluminescence sandwich immunoassay. Briefly, in the first incubation a biotinylated monoclonal anti-TNNT2-specific antibody and a monoclonal anti-TNNT2-specific antibody labeled with ruthenium react to form a sandwich complex (the antibodies recognize two epitopes: amino acid positions 125–131 and 136–147 of the 288 TNNT2 amino acids). During the second incubation, after addition of streptavidin-coated microparticles, the complex becomes bound to the solid phase via interaction of biotin and streptavidin. Analyte concentration is automatically calculated via a calibration curve in ng/mL. Internal quality controls are run every 8 hours (IQC material – Technopath Multichem 3 levels). The assay is calibrated monthly as required for IQC drift. Recommended 99th percentile cut-offs are14.0 ng/liter for the whole population with a CV of less than 10% (9.0 ng/liter for women; 16.8 ng/liter for men).

**GPNMB Immunoassay validation**

GPNMB concentration was quantified in a solid phase sandwich ELISA using the R&D Systems Human Osteoactivin/GPNMB DuoSet ELISA kit (Catalog number: DY2550).  This clinically validated method utilizes a sandwich ELISA performed on a microtiter plate coated with a goat anti-human GPNMB capture antibody. Standards and samples are added to the plate and incubated, allowing GPNMB to bind. Detection is achieved using a biotinylated goat anti-human GPNMB antibody, followed by Streptavidin-HRP. A chromogenic substrate is then added, generating a colorimetric signal proportional to the bound

GPNMB. Samples are measured in duplicate, and results are expressed in pg/mL. For detailed protocol please see the kit manufacturer insert.

**References**


1. Kauffmann A, Gentleman R, Huber W. arrayQualityMetrics--a bioconductor package for quality assessment of microarray data. Bioinformatics 2009;25:415-416.
2. Searle S, Speed F, Milliken G. Population marginal means in the linear model: an alternative to least square means. The American Statistician 2012;34:216-221.
3. Kuznetsova A, Brockhoff PB, Christensen RHB. lmerTest Package: Tests in Linear Mixed Effects Models. Journal of Statistical Software 2017;82:1 - 26.
4. Ritchie ME, Phipson B, Wu D, et al. limma powers differential expression analyses for RNA-sequencing and microarray studies. Nucleic acids research 2015;43:e47.
5. Yanai I, Benjamin H, Shmoish M, et al. Genome-wide midrange transcription profiles reveal expression level relationships in human tissue specification. Bioinformatics 2005;21:650-659.
6. Subramanian A, Tamayo P, Mootha VK, et al. Gene set enrichment analysis: a knowledge-based approach for interpreting genome-wide expression profiles. Proceedings of the National Academy of Sciences of the United States of America 2005;102:15545-15550.
7. Korotkevich G, Sukhov V, Budin N, Shpak B, Artyomov MN, Sergushichev A. Fast gene set enrichment analysis. bioRxiv 2021:060012.
