## Supplementary material for "Skeletal muscle biomarkers of amyotrophic lateral sclerosis: a large-scale, multi-cohort proteomic study": eTable 1

**eTable 1. Participant Characteristics (CSF)**

| | | Discovery Cohort | | Replication Cohort | |
|---|---|---|---|---|---|
| | | **ALS** [a] | **Controls** | **ALS** [b] | **Controls** |
| **Study Cohort** | | | | | |
| Number of participants | N | 70 | 51 | 82 | 25 |
| Baseline age, years | Median (Q1-Q3) | 58 (51-67) | 47 (37-58) | 58 (50-66) | 57 (33-60) |
| Sex, male | N (%) | 41 (59%) | 23 (45%) | 45 (55%) | 17 (68%) |
| Baseline ALSFRS-R | Median (Q1-Q3) | 39 (34-44) | NA | 36 (31-41) | NA |
| Months since onset | Median (Q1-Q3) | 12.9 (4.0-27.2) | NA | 27.4 (16.3-37.8) | NA |
| Baseline ΔFRS, point(s)/month | Median (Q1-Q3) | 0.7 (0.3-1.1) | NA | 0.4 (0-2.8) | NA |
| **Longitudinal Subset** | | | | | |
| Number of participants | N | 30 | 24 | 40 | 0 |
| # of visits per person | Median (range) | 4 (2-11) | 3 (2-5) | 3 (2-3) | 1 |

ALSFRS-R = ALS Functional Rating Scale, Revised; ΔFRS = deltaFRS, calculated as (48 - baseline ALSFRS-R) / (months from onset to baseline); Q1 = 1st quartile; Q3 = 3rd quartile.

[a] Includes 21 *SOD1*, 12 *C9orf72*, 2 other genes, and 35 with no pathogenic variant identified.

[b] Includes 3 *C9orf72*.
