## Supplementary material for "Skeletal muscle biomarkers of amyotrophic lateral sclerosis: a large-scale, multi-cohort proteomic study": eTable 2

**eTable 2. Biomarker Contexts of Use**

| Biomarker | Full Name | Context of Use | | | | Notes |
|---|---|---|---|---|---|---|
| | | Disease State: ALS (v. Ctrl) [a] | In Clinically Manifest ALS | | | |
| | | | Prognosis: Worse (v. Better) [b] | Monitoring | | |
| | | | | Disease progression [c] | Temporally stable [d] | |
| ANTXR2 | Anthrax toxin receptor 2 | − (P) | | ↓ (P) | | Schwann cells, ECM interaction [1] |
| ART3 | ADP-ribosyltransferase 3 | − (P) | | ↓ (P) | | Skeletal muscle |
| PDLIM3 | PDZ and LIM Domain 3 | + (P,C) | + (C) | ↑ (P) | | Skeletal muscle, ECM interaction [2] |
| MYL11 | Myosin regulatory light chain 11 | + (P,C) | + (C) | ↑ (P) | | Skeletal and cardiac muscle |
| CA3 | Carbonic anhydrase 3 | + (P,C) | | | | Skeletal muscle |
| APOBEC2 | Apolipoprotein B mRNA editing enzyme catalytic subunit 2 | + (P) | | | | Skeletal muscle |
| TNNT2 | Cardiac Troponin T2 | + (P) | | ↑ (P) | | Skeletal muscle |
| TNNI2 | Troponin I2, fast skeletal muscle | + (P) | | | | Skeletal muscle |
| ACTN2 | Alpha-actinin 2 | + (P) | | | + stable (P) | Skeletal and cardiac muscle |
| MYOM2 | Myomesin 2 | + (P) | | | + stable (P) | Skeletal muscle, NK cells |
| CAPN3 | Calpain-3 | + (P) | | | + stable (P) | Skeletal muscle, oligodendrocytes |
| KLHL41 | Kelch-like protein 41 | + (P) | | | + stable (P) | Skeletal muscle |
| HSBP6 | Heat shock protein β6 | + (P) | | | | Skeletal muscle |
| NEFH | Neurofilament heavy chain | + (C) | | | | Central and peripheral axons |
| NEFL | Neurofilament light chain | + (C) | + (C) | | | Central and peripheral axons |
| CHIT1 | Chitinase 1 | + (C) | + (C) | | | Macrophages |
| GPNMB | Glycoprotein non-metastatic melanoma protein B | + (C) | + (C) | | | Macrophages and microglia |
| GFAP | Glial fibrillary acidic protein | + (C) | + (C) | | | Astrocytes |
| PLIN3 | Perilipin 3 | | + (P) | | | Skeletal muscle lipid droplet protein [3] |
| CRTAC1 | Cartilage acidic protein 1 | | − (P) | | | ECM |
| CLEC3B | C-type lectin domain family member B | | − (P) | ↓ (P) | | Microglia |
| ANTXR1 | Anthrax toxin receptor 1 | | | ↓ (P) | | Astrocytes |
| GDF11/MSTN | Growth differentiation factor 11/myostatin | | | ↓ (P) | | Skeletal muscle |
| ITGAV/ITGB3 | Integrin alpha-V/beta-3 | | | ↓ (P) | | ECM, microglia |
| EHMT2 | Euchromatic histone lysine methyltransferase 2 | | | ↓ (P) | | Brain and peripheral nerve |
| PTN | Pleiotrophin | | | ↑ (P) | | Brain |

(P) = In plasma. (C) = In CSF. (P,C) = In plasma and CSF. Ctrl = Control. ECM = Extracellular matrix.

[a] "−" denotes lower in ALS compared to controls. "+" denotes higher in ALS compared to controls.

[b] "−" denotes lower level associated with worse prognosis. "+" denotes higher level associated with worse prognosis.

[c] "↓" denotes decreasing over time. "↑" denotes increasing over time.

[d] "+ stable" denotes elevated in ALS compared to controls, but stable over time.
