## Supplementary figures and images for "Skeletal muscle biomarkers of amyotrophic lateral sclerosis: a large-scale, multi-cohort proteomic study"

### eFigure 1

#### eFigures

##### eFigure 1. Study Schema

**eFigure 1. Study Schema**

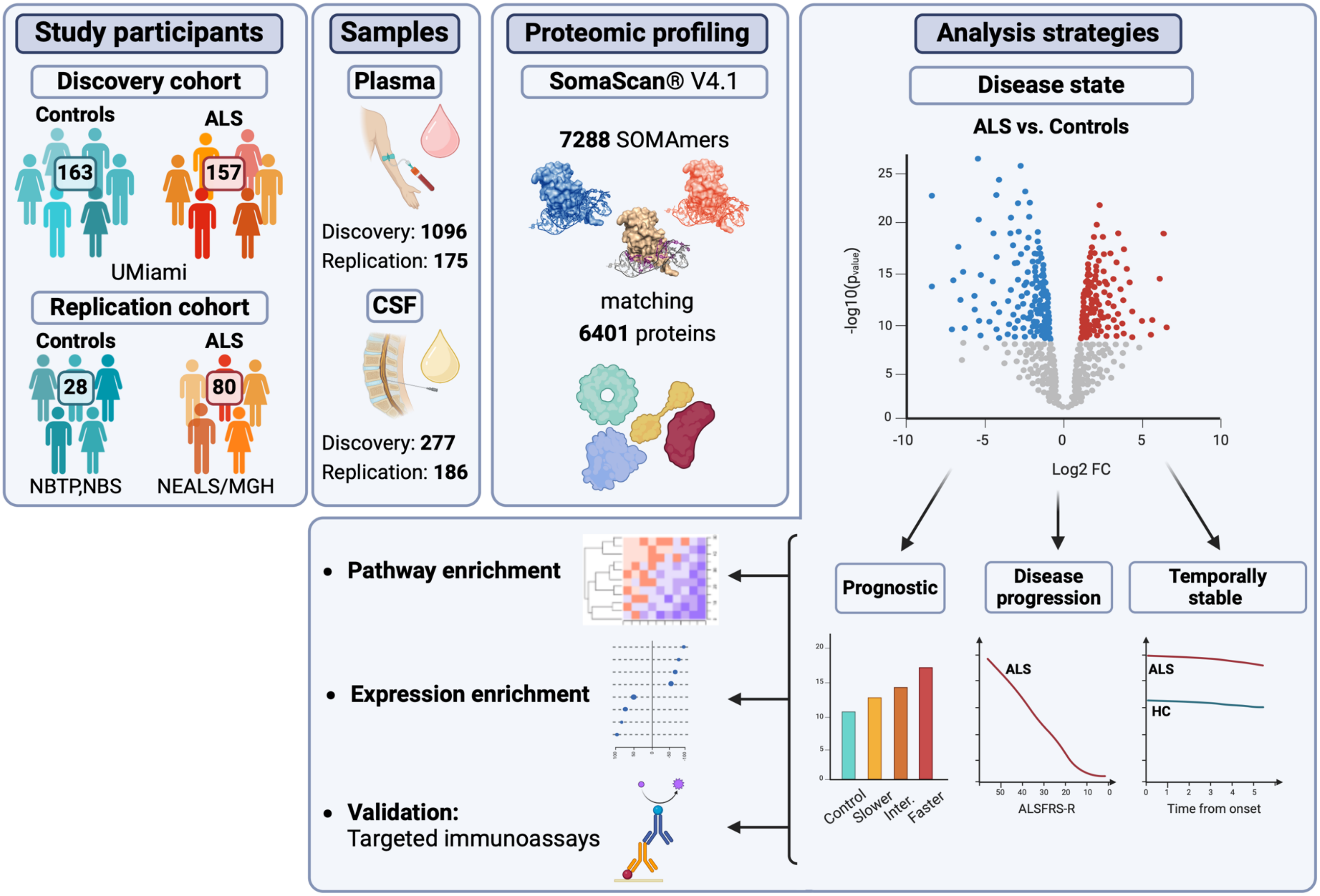
