## Supplementary material for "Skeletal muscle biomarkers of amyotrophic lateral sclerosis: a large-scale, multi-cohort proteomic study": eFigure 2

**eFigure 2**. **Principal Component Analysis**

Principal component analysis (PCA) showed no major batch effect, but still significant associations between key clinical parameters, plateID and hemoglobin levels with the top 10 principal components (PCs). Heatmaps summarize the level of significance of the associations, in plasma **(A)** and CSF **(C)**. Scatterplots show combination of the first two PCs by clinical parameters and plate ID, in plasma **(B)** and CSF **(D)**. In the heatmaps, adjusted p-values <0.001 denoted by ***; 0.001-0.01 denoted by **; 0.01-0.05 denoted by *; and 0.05-0.1 by “x”. Affected = Disease state (yes for ALS, no for control). CollAge = Age at sample collection. PlateId = Plate ID.

### eFigure 2. Principal Component Analysis

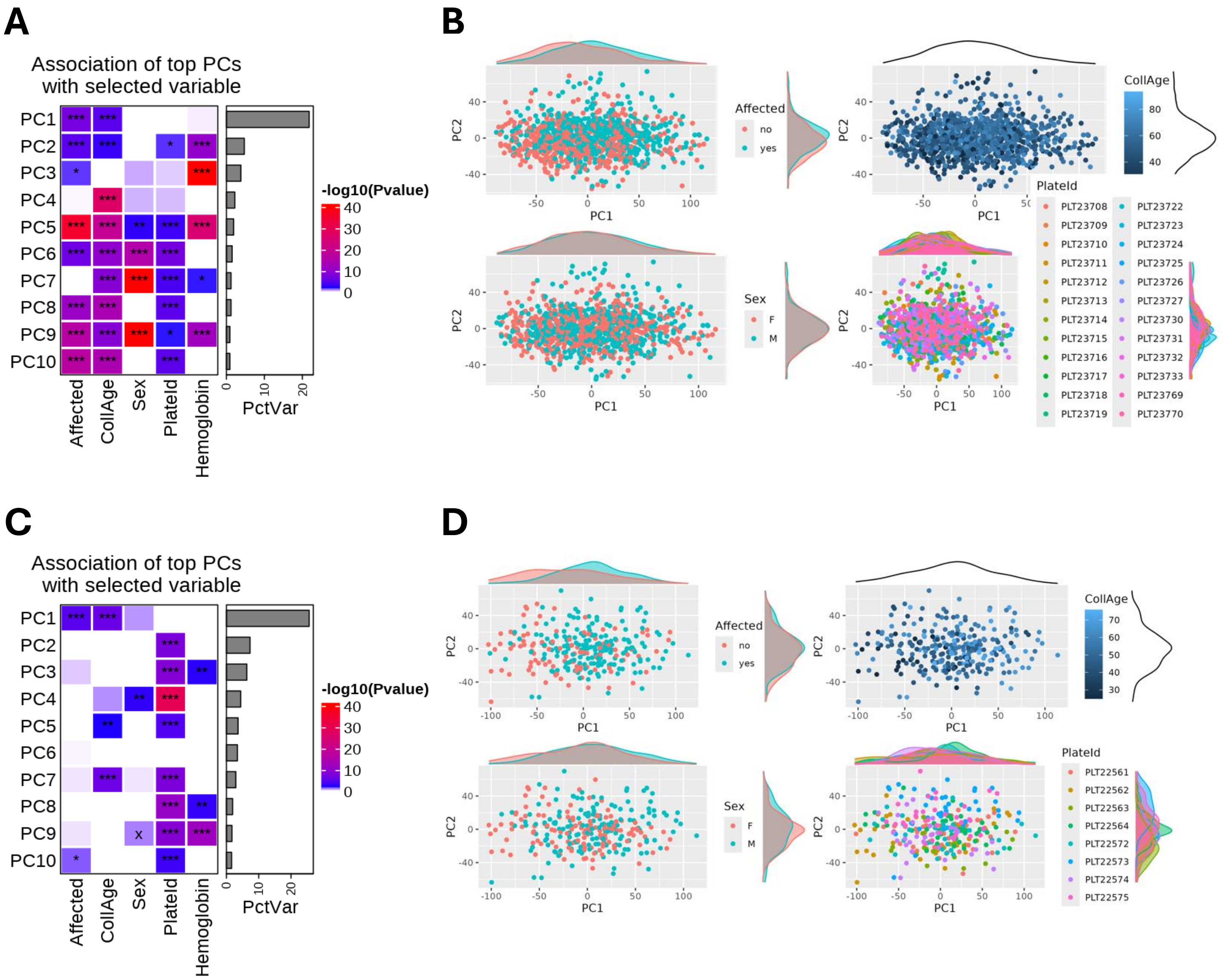
