## Supplementary material for "Skeletal muscle biomarkers of amyotrophic lateral sclerosis: a large-scale, multi-cohort proteomic study": eFigure 3

**eFigure 3. Consistency of Biomarker Candidates Identified by Different Statistical Methods**

Heatmap showing top 10 proteins with lowest adjusted p-values when comparing ALS to controls, in CSF and plasma, as identified by linear mixed effect models (LME) run on all samples; limma modeling performed on all samples; and limma modeling performed on baseline samples only. Adjusted P-value <0.001 denoted by ***; 0.001-0.01 denoted by **; 0.01-0.05 denoted by *; and 0.05-0.1 by "." (period).

### eFigure 3. Additional CSF Markers Differentially Expressed in ALS vs. Healthy Controls

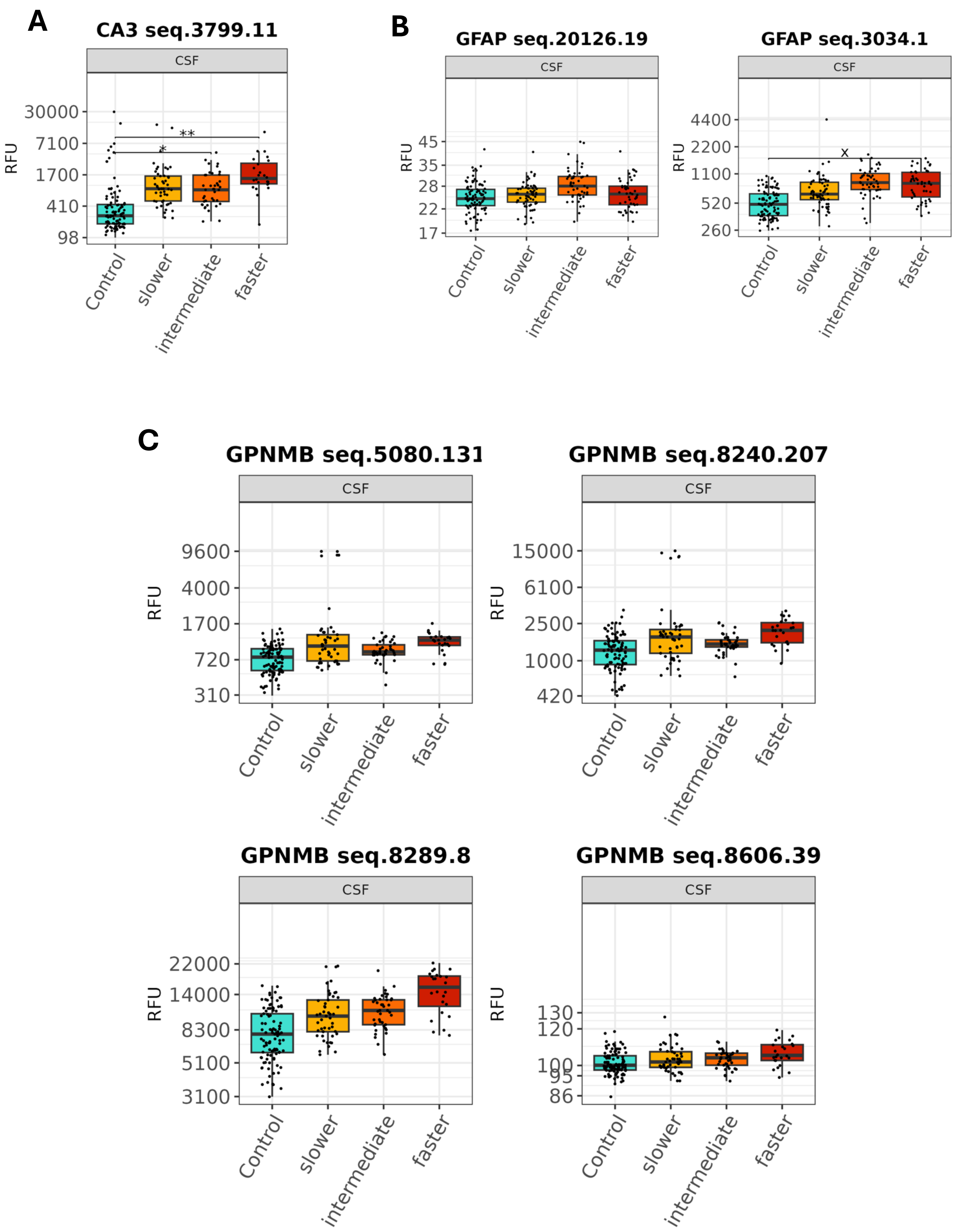
