## Supplementary material for "Skeletal muscle biomarkers of amyotrophic lateral sclerosis: a large-scale, multi-cohort proteomic study": eFigure 4

**eFigure 4. Changes in Plasma GDF11|MSTN Are Driven by MSTN**

Boxplots showing normalized age-, sex- and plate-adjusted SOMAmer intensities for GDF11 and/or MSTN in ALS vs. controls, and accompanying scatterplots to illustrate the association between GDF11/MSTN and ALSFRS-R in ALS. (**A**) GDF11|MSTN SOMAmer (seq 2765.4) is significantly lower in ALS compared to controls, and it declines with falling ALSFRS-R (adjusted p-value [padj] < 0.05). (**B**) MSTN SOMAmer (seq 12077.32) is also significantly lower in ALS compared to controls, and declines with falling ALSFRS-R (adjusted p-value < 0.05). (**C**) MSTN SOMAmer (seq 14583.49) levels, however, are comparable between ALS and controls, and relatively stable in ALS (i.e. not associated with ALSFRS-R, adjusted p-value > 0.05). (**D**) Similarly, GDF11 SOMAmer (seq 14587.16) levels are comparable between ALS and controls, and relatively stable in ALS. *** denotes p-value<0.001. "ns" denotes not statistically significant (p-value>0.1).

**eFigure 4 . Consistency of Biomarker Candidates identified by Different Statistical Methods**

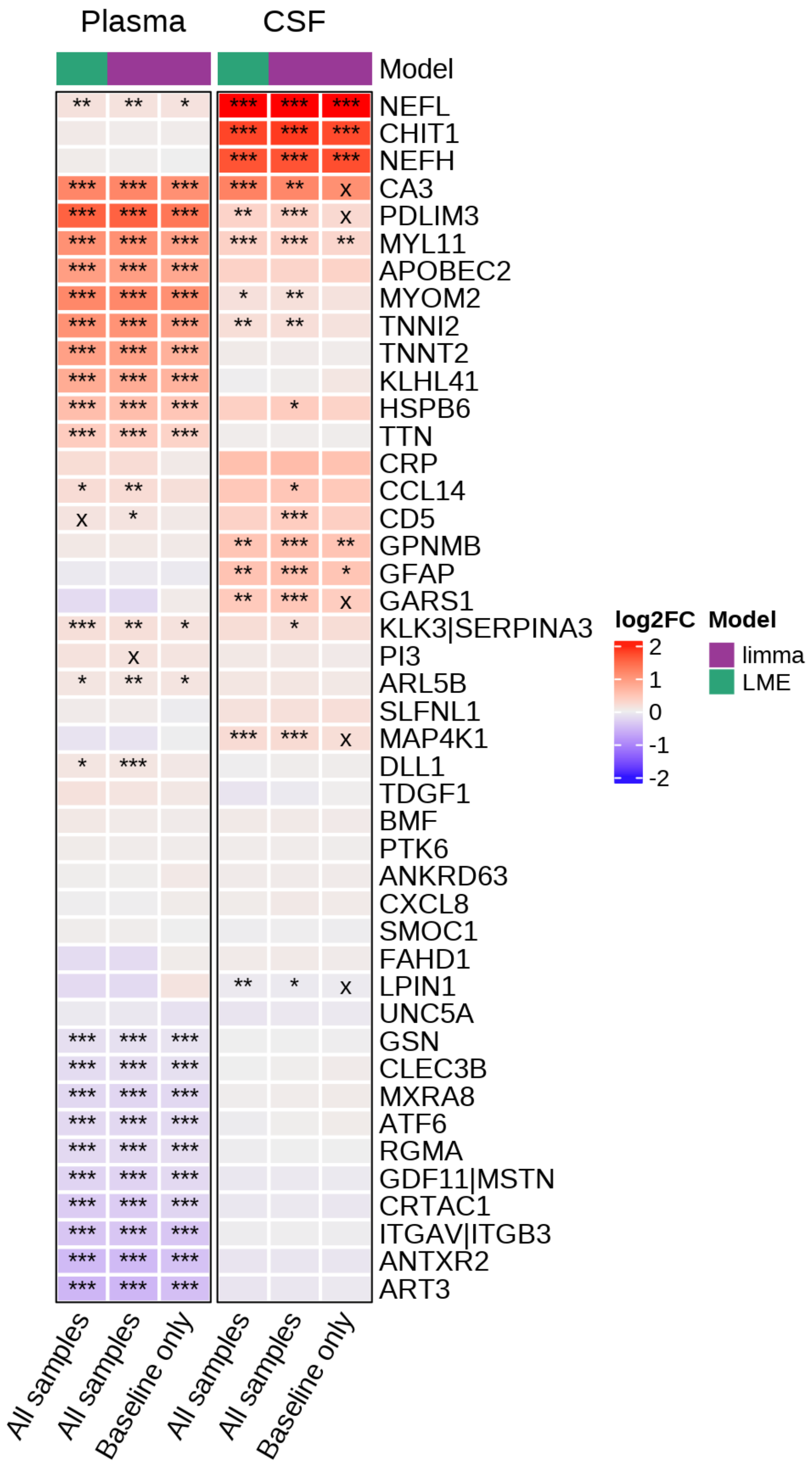
