## Supplementary material for "Skeletal muscle biomarkers of amyotrophic lateral sclerosis: a large-scale, multi-cohort proteomic study": eFigure 5

**eFigure 5. Plasma Biomarkers of Disease Progression**

Volcano plot of longitudinal change in SOMAmer intensity per 1 year follow-up from baseline. The intensity of SOMAmers to the left (e.g. ANTXR2, ART3) fell with increasing follow-up (and disease) duration. The intensity of SOMAmers to the right (e.g. TNNT2, PTN, PDLIM3) increased with longer follow-up (and disease) duration. SOMAmers marked in red are significant, with an adjusted p-value of <0.05, estimated by linear mixed model fitted to longitudinal subset of ALS cases.

**eFigure 5. Changes in Plasma GDF11|MSTN Are Driven by MSTN**

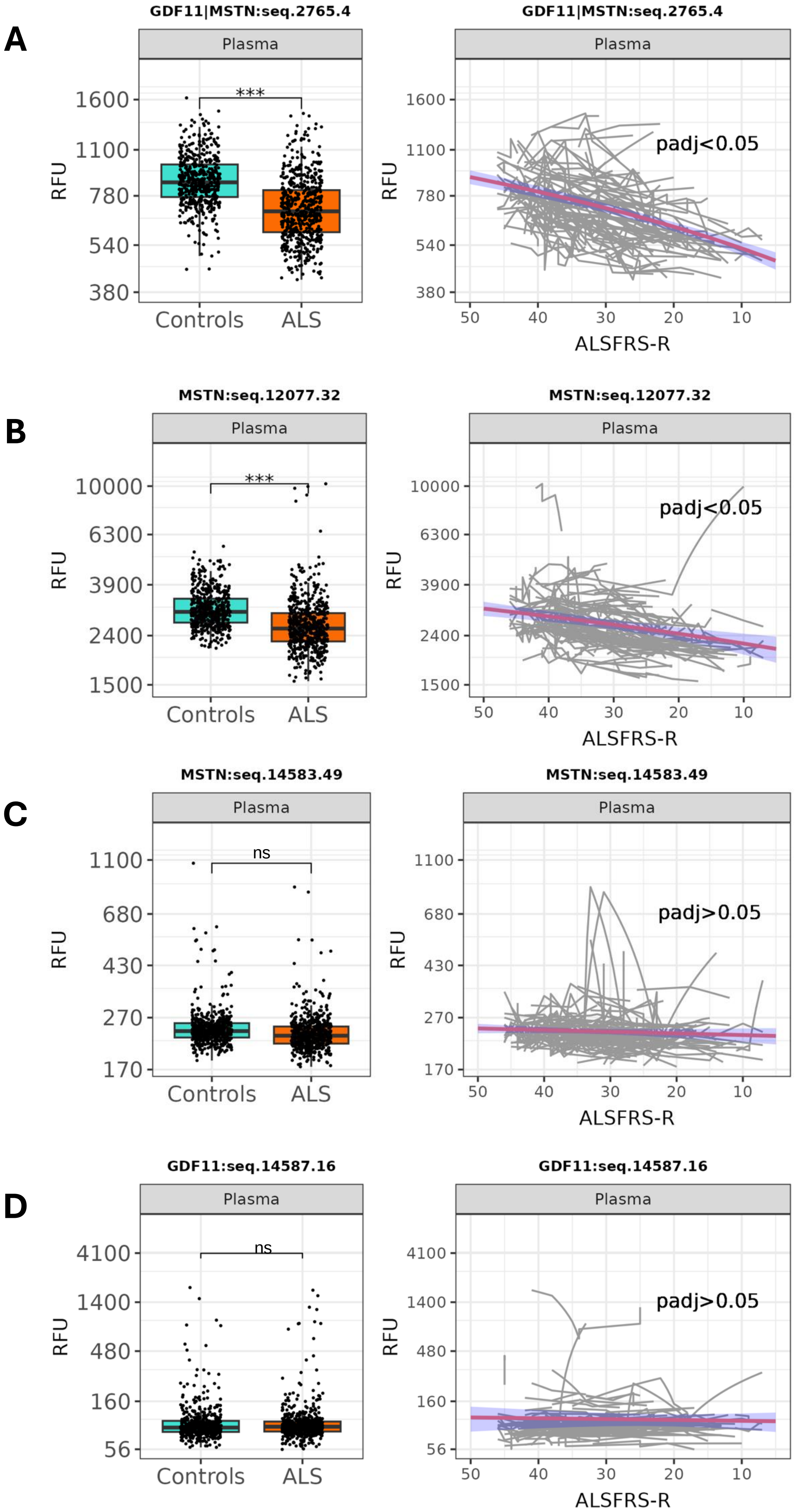
