## Supplementary material for "Skeletal muscle biomarkers of amyotrophic lateral sclerosis: a large-scale, multi-cohort proteomic study": eFigure 6

**eFigure 6. Additional CSF Markers Differentially Expressed in ALS vs. Healthy Controls**

In addition to those shown in eFigures 1F-1I, boxplots illustrating normalized age-, sex-, and plate-adjusted SOMAmer intensities for CA3 (seq 3799.11) **(A)**, GFAP (seq 20126.19, 3034.1) **(B)**, and GPNMB (seq 5080.131, 8240.207, 8289.8, 8606.39) **(C),** in controls and in ALS subgroups based on estimated progression rate. Adjusted P-values = 0.001-0.01 denoted by **; 0.01-0.05 denoted by *; and 0.05-0.1 by “x”.  RFU = relative fluorescence units.

**eFigure 6. Plasma Biomarkers of Disease Progression**

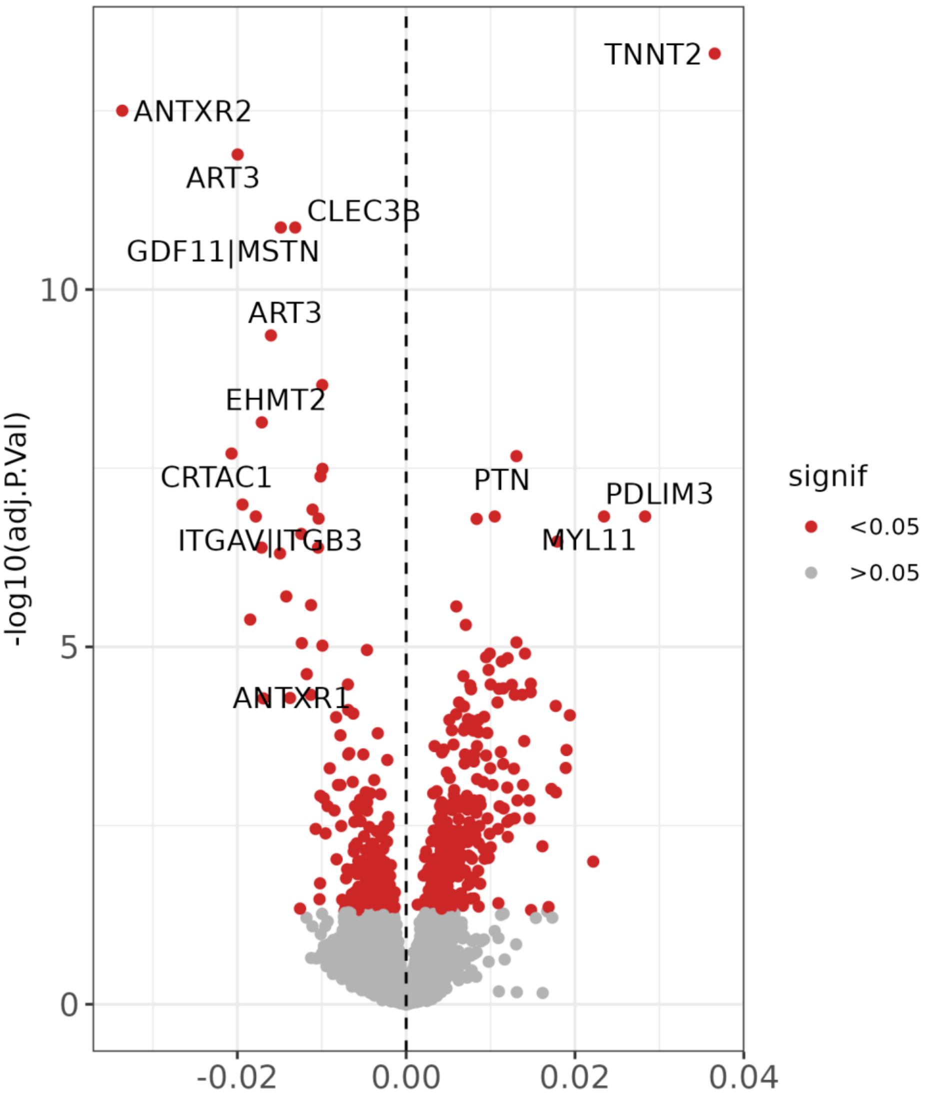


Coefficient: change of protein level on log2 scale
per 1 month of follow up since baseline
