## Supplementary material for "Skeletal muscle biomarkers of amyotrophic lateral sclerosis: a large-scale, multi-cohort proteomic study": eFigure 7

**eFigure 7. Biomarker Candidates Replicated in an Independent Cohort**

Heatmap comparing plasma (**A**) and CSF (**B**) results from the discovery cohort vs. replication cohort. Top 10 SOMAmers with lowest adjusted p-value from two analyses – comparison of ALS vs. healthy control (left panel) and association with ALSFRS-R (right panel) – from each cohort are included here. In **A** and **B**, left panels show the log2 fold-change, where positive (or negative) values signify higher (or lower) intensities in ALS compared to controls; right panels show the change of SOMAmer intensities (on a log2 scale) per 1 point change in ALSFRS-R in the ALS group, where a negative (or positive) association, represented by the coefficient (Coef), indicates that the protein level rises (or falls) as ALSFRS-R decreases with disease progression. P-value <0.001 denoted by ***; 0.001-0.01 denoted by **; 0.01-0.05 denoted by *; and 0.05-0.1 by "x".

**eFigure 7. Biomarker Candidates Replicated in an Independent Cohort**

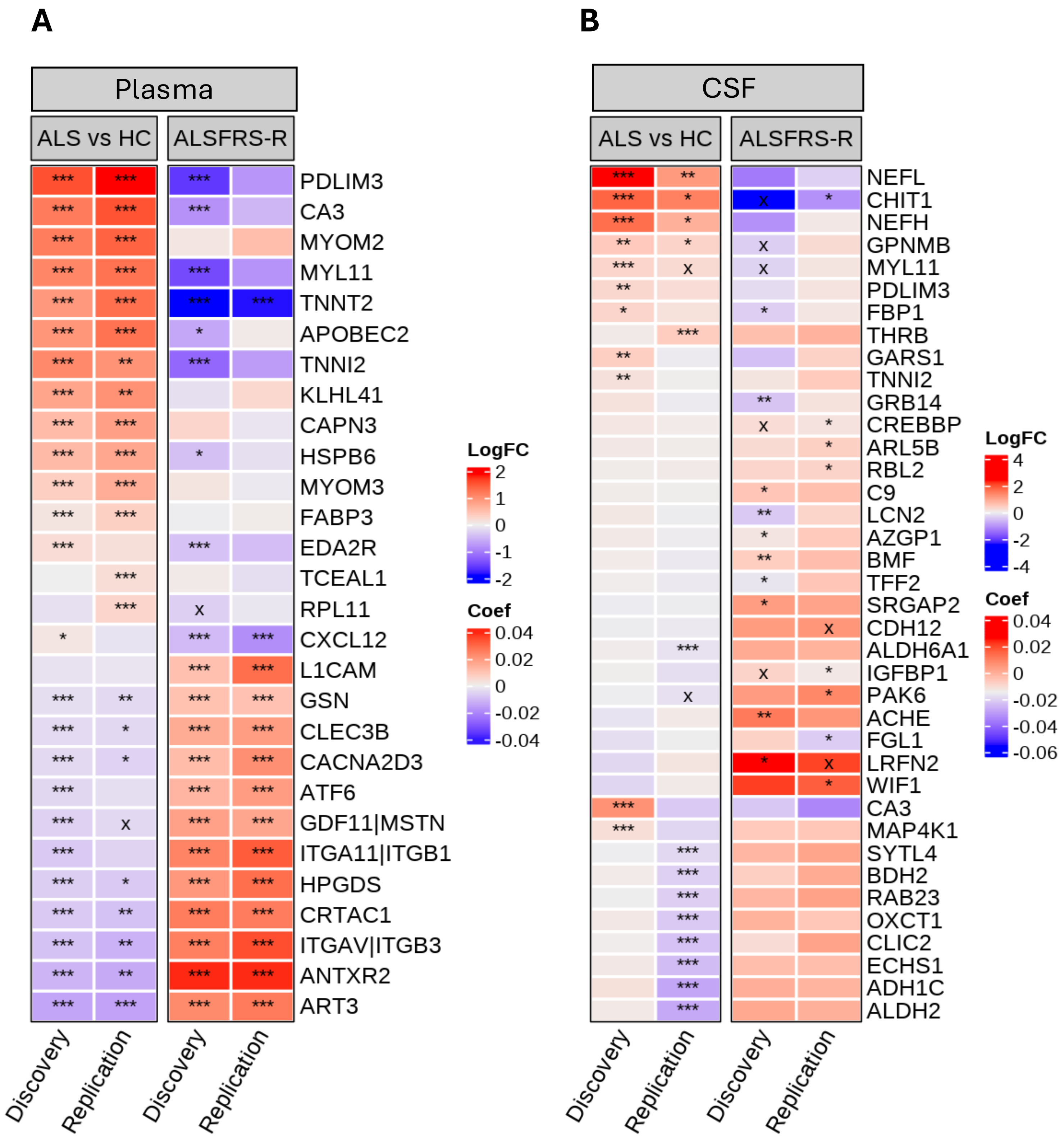
