## Supplementary material for "Skeletal muscle biomarkers of amyotrophic lateral sclerosis: a large-scale, multi-cohort proteomic study": eFigure 8

**eFigure 8. Plasma TNNT2 Immunoassay Confirmation**

(**A**) Scatterplot showing the correlation between TNNT2 SOMAmer intensity and TNN2 concentration quantified using the Roche Elecsys immunoassay. (**B**) Boxplots showing ELISA concentrations and SOMAmer intensity in ALS and controls; data are derived from linear effects model adjusting for age, sex and within-person correlation. *** indicates p-value < 0.001. (**C**) Scatterplot showing the increase in TNNT2 (measure by ELISA) as a function of time since baseline among ALS participants with longitudinal evaluations and TNNT2 immunoassay measurements.

**eFigure 8. Plasma TNNT2 Immunoassay Confirmation**

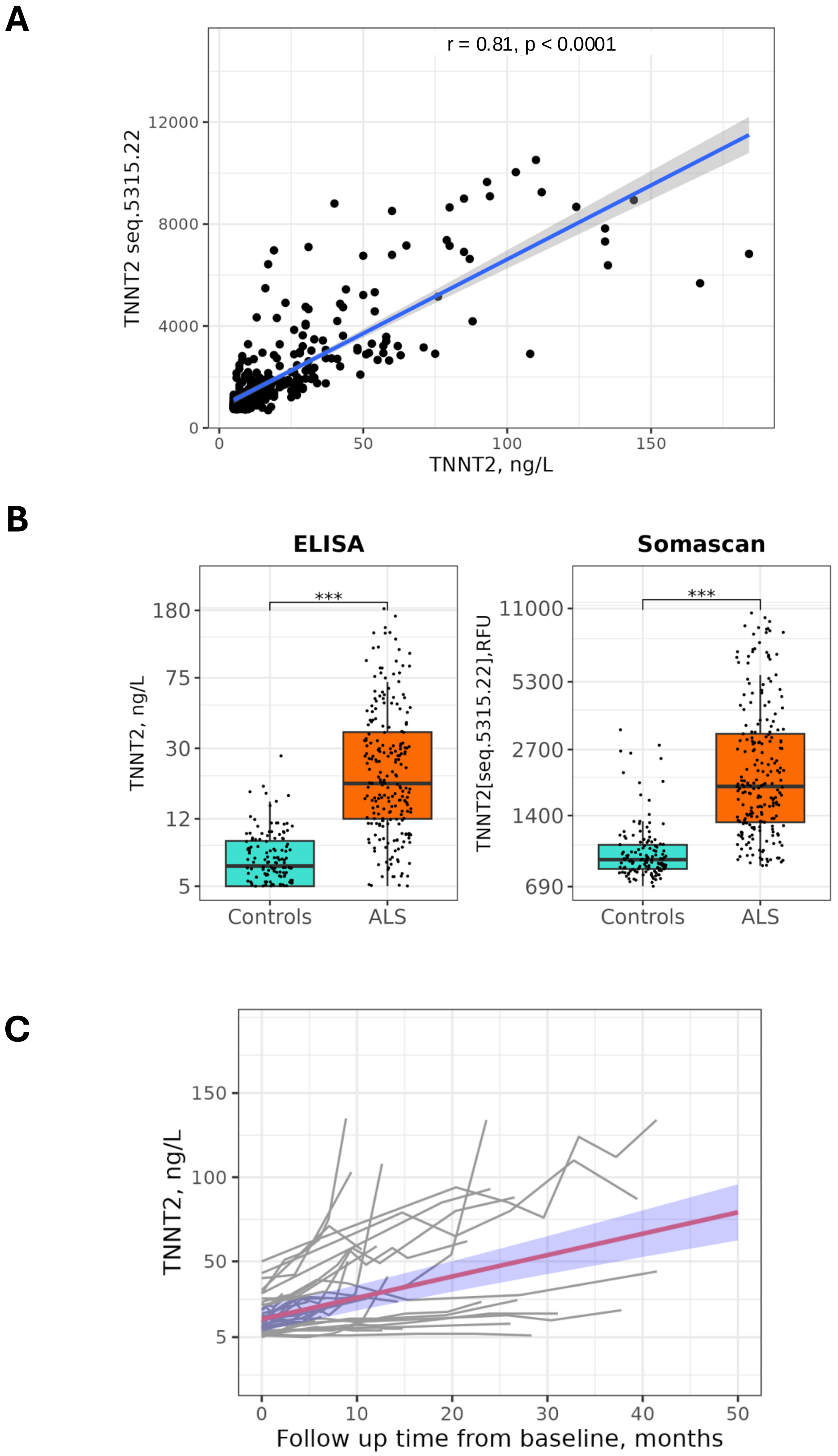
