## Supplementary material for "Skeletal muscle biomarkers of amyotrophic lateral sclerosis: a large-scale, multi-cohort proteomic study": eFigure 9

**eFigure 9. CSF GPNMB Immunoassay Confirmation**

**(A)** Scatterplot showing the correlation between GPMNB SOMAmer intensity and GPNMB concentration quantified using the R&D Systems Human Osteoactivin/GPNMB DuoSet immunoassay. **(B)** Boxplots showing ELISA concentrations and SOMAmer intensity in ALS and controls; data are derived from linear effects model adjusting for age, sex and within-person correlation. ** indicates p-value < 0.01.

**eFigure 9. CSF GPNMB Immunoassay Confirmation**

**A**

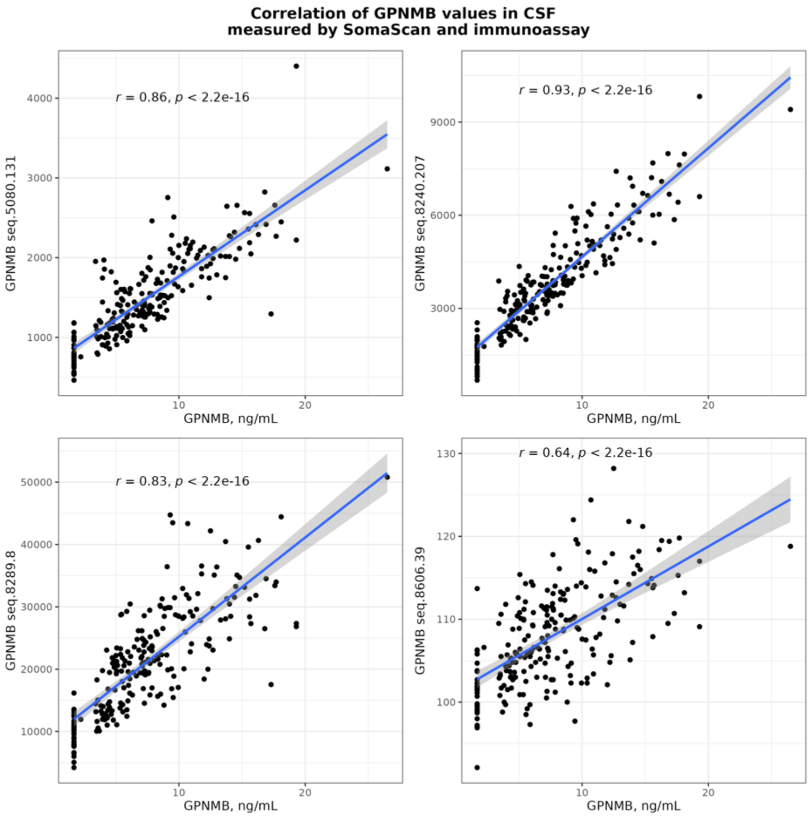


**B**

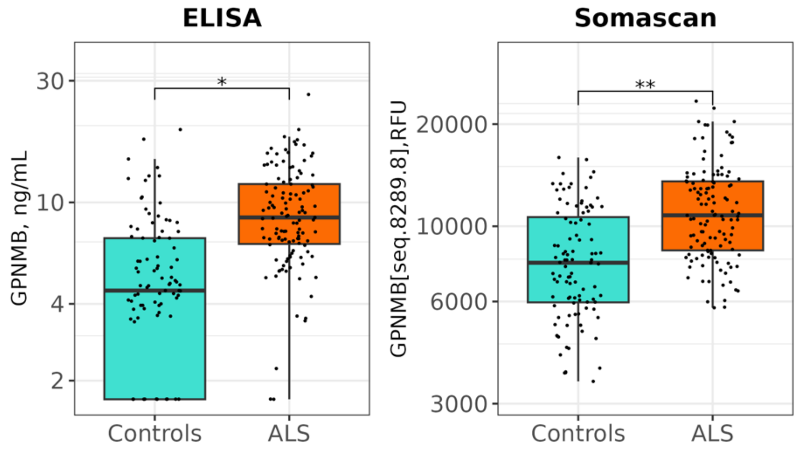
