## Supplementary material for "Skeletal muscle biomarkers of amyotrophic lateral sclerosis: a large-scale, multi-cohort proteomic study": eFigure 10

**eFigure 10. Tissue Expression-Based Enrichment by Context of Use**

Scatterplots showing the relationship between median tau parameter (tissue specificity) and median log2FC for proteins identified as significantly differentially regulated (adjusted p-value < 0.05) in **(A)** ALS vs control, and **(B)** temporally stable biomarkers. **(C)** Scatterplot for disease progression biomarkers, showing the relationship between median tau parameter and median log2FC in protein level per one point change in ALSFRS-R.

**eFigure 9. Tissue Expression-Based Enrichment by Context of Use**

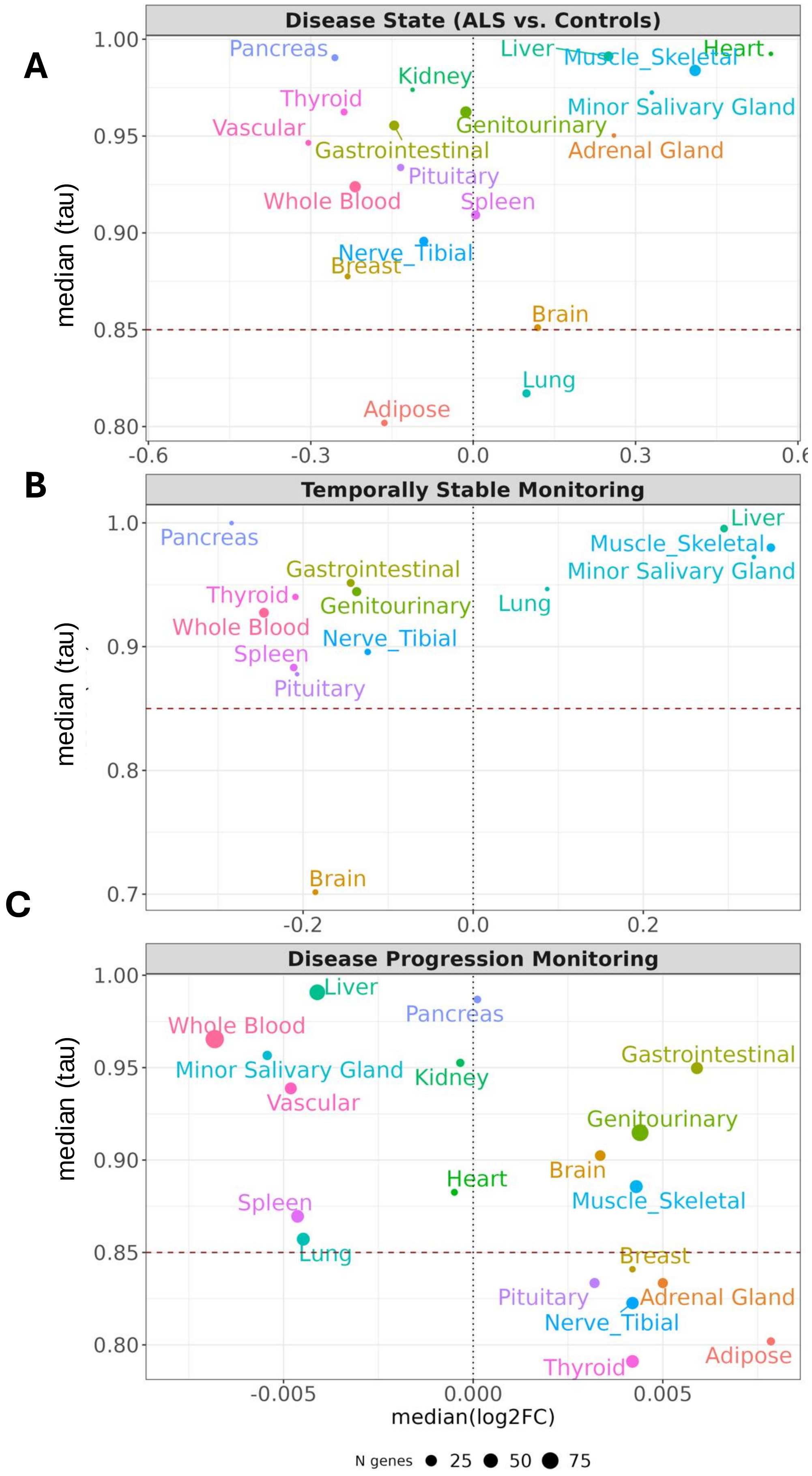
